# Monitoring-based and self-reported close-contact records in relation to ultra-wideband-derived proximity in a long-term care facility: a single-facility observational study

**DOI:** 10.64898/2026.04.10.26350570

**Authors:** Hayato Shinto, Gerardo Chowell, Yoshihiro Takayama, Yu Ohki, Kei Saito, Kenji Mizumoto

## Abstract

**Background:** In long-term care facilities (LTCFs), close-contact identification often relies on staff recall and monitoring records because residents may be unable to self-report reliably. How these different record-generation processes relate to proximity-based sensor measurements in routine LTCF workflow remain unclear, and how such differences may influence contact-based decision-making in outbreak response is not well understood.

**Methods:** We conducted a five-day observational study in a Japanese LTCF using ultra-wideband (UWB) indoor positioning. Twenty-seven participants wore UWB tags, including 16 residents and 11 staff members; 10 staff members completed questionnaires. We compared UWB-derived proximity with questionnaire-derived contacts from staff self-report and monitoring-based proxy records, and assessed directional discrepancies under multiple distance–time thresholds.

**Results:** Questionnaire-based records and UWB-derived proximity showed different patterns of discrepancy across contact types. Within this facility, resident-related monitoring-based proxy records showed relatively small directional discrepancies, whereas staff self-reports tended to identify additional resident-staff contacts under the baseline threshold (≤1.0 m for ≥15 min). Several alternative thresholds were associated with discrepancies closer to zero than the baseline, although the apparent ranking varied by summary metric.

**Conclusions:** In this single-facility observational study, different contact-list generation processes were associated with different patterns of discrepancy relative to a proximity-based operational measure. These findings support interpretation in terms of workflow-specific contact-list generation rather than a single universally optimal threshold and may help inform facility-level review of contact identification practices in LTCFs. These findings support aligning contact identification strategies with facility-specific workflows to improve the feasibility and effectiveness of IPC practices in LTCFs.

## Introduction

Long-term care facilities (LTCFs) are repeatedly affected by outbreaks of respiratory and gastrointestinal pathogens because they house older adults with multiple comorbidities and require frequent close-range caregiving in predominantly indoor, shared environments. When outbreaks occur, timely identification of close contacts underpins core public health actions and operational decisions—including targeted testing, isolation, cohorting, and staff work restrictions—and therefore influences both outbreak control and the continuity of care [1].

In many LTCFs, close contacts are identified through interviews with staff and review of care records (e.g. assignment rosters, activity logs). However, the completeness and consistency of these approaches can be limited: residents may be unable to report contacts due to cognitive impairment, and staff must reconstruct numerous brief encounters under time pressure. As a result, operational definitions based on distance–time thresholds (commonly ‘within 1–2 metres for 10–15 minutes’) may be applied inconsistently across individuals and shifts, leading to both under-identification (risking missed transmission chains) and over-identification (increasing unnecessary testing, isolation, and staff shortages) [2–5]. These challenges highlight the need to better understand how different contact identification approaches function within routine LTCF workflows.

Wearable proximity sensing technologies can capture contacts without relying solely on retrospective reporting and offer an opportunity to validate and refine LTCF contact tracing protocols [6, 7]. Among available systems, ultra-wideband (UWB) indoor positioning provides high spatial resolution that can quantify second-by-second interpersonal distance [8, 9]. By linking objective proximity measurements to staff-reported contact lists, UWB data could help identify systematic reporting biases and support setting-appropriate, transparent thresholds for action during outbreak investigations. However, proximity-based measures capture physical distance rather than infection risk directly, and their interpretation in real-world IPC workflows requires careful evaluation. Previous work in long-term institutional settings has shown heterogeneity in contact patterns by ward and staff category [11], while recent reviews and care-home feasibility studies suggest that contact-tracing technologies remain methodologically immature and operationally challenging in practice [12,13]. Nevertheless, evidence from real-world LTCFs directly comparing staff-generated contact records with high-resolution UWB-measured proximity remains scarce.

Here, we conducted a five-day observational study in a Japanese LTCF to compare staff self-report and monitoring-based proxy records in relation to UWB-derived proximity. We assessed discrepancies by contact type and explored alternative distance–time thresholds. We framed the study as an operational comparison of contact-list generation workflows within a single facility, rather than as an evaluation of a transmission gold standard or a population-level estimate of contact behaviour.

## Methods

### Study design and setting

This observational study was conducted in December 2024 at an LTCF in Kyoto Prefecture, Japan. The facility has multiple floors; two adjacent care areas on one floor were included in this study, and staff may move between them as part of routine care. Residents spend their daily lives in their own rooms and shared spaces; each area is equipped with a bathroom and toilet. Each area houses 8-10 residents, and one to two staff members are assigned to each area 24 hours a day on a rotating basis. The survey was conducted over five consecutive days, from Wednesday to Sunday. The experiment began at 9:30 AM on Wednesday and ended at 11:59 PM on Sunday.

Because staff worked in shifts, they wore UWB tags only during working hours. Staff working periods used in the analyses were determined from tag-wearing records rather than from questionnaire responses. Residents wore UWB tags continuously and therefore replaced tags every one to two days due to battery life. A total of 42 receivers were installed across the two study areas. Measurement conditions differed between participant groups because of operational constraints in this facility and should be considered when interpreting discrepancies between reporting methods and

UWB-derived proximity. This study was designed as an operational comparison within a single LTCF and was not intended to estimate generalisable performance characteristics of contact identification across facilities.

### Participants

Of 29 eligible individuals, 27 (16 residents and 11 staff; 93.1%) participated and wore UWB tags. Written informed consent was obtained from facility staff, while residents were included using an opt-out approach. Questionnaire responses were obtained from 10 facility staff; analyses involving questionnaires were restricted to these respondents and the 16 residents for whom staff reported monitoring information.

### Questionnaire survey

At the end of the survey period, participating staff completed a structured questionnaire (Additional file 2) developed for this study. Items captured staff role/assignment, and daily lists of close contacts. For the purpose of the questionnaire, a close contact was defined as a person who was within 1 metre for a cumulative total of ≥15 minutes during a day, regardless of mask use. Because residents were not surveyed, staff additionally reported residents’ close contacts, and these monitoring reports were combined into questionnaire-based resident contact data. Although the questionnaire form asked staff to report contacts up to before 9:00 AM on Monday, December 16, responses referring to that period were excluded from the comparative analyses so that questionnaire-based contact data corresponded to the analysed UWB period through 11:59 PM on Sunday.

### UWB positioning measurements

The UWB indoor positioning system comprised wearable transmitters (tags) and fixed receivers. The system captured location data at 1 Hz (once every second) with a nominal measurement error of 10-30 cm [8, 9]. However, because residents were asked to wear tags continuously for 24 hours and study personnel were not permitted to enter the facility between 20:00 and 07:00, battery replacement could not be performed overnight. To maintain continuous recording throughout the observation period, staff tags were recorded at 1 Hz (once every second), whereas resident tags were recorded once every 10 seconds. Forty-two receivers were deployed across the two study areas. Tags were attached to the participant’s chest to approximate torso position during interaction when feasible (typically for staff). For residents, long-term chest placement was not always feasible; tags were therefore attached to alternative locations such as an arm band or a wheelchair, depending on the individual. These differences in sampling frequency and tag placement reflected operational constraints in this facility and should be considered when interpreting resident-related and staff-related discrepancies relative to UWB-derived proximity. We note that UWB-derived proximity was treated as an operational measure of interpersonal distance rather than a direct measure of infection risk.

### Data processing: raw data preparation

The raw data were trimmed to cover the period from 9:30 AM on Wednesday to 11:59 PM on Sunday. Tag IDs were then matched to participant IDs.

### Smoothing and interpolation

To improve positional accuracy, we applied Kalman filtering and a Long Short-Term Memory (LSTM) model, both of which have been validated in previous UWB positioning studies [10]. The LSTM model used a rolling input of the preceding 120 seconds to interpolate missing points and smooth coordinate fluctuations. Unlike prior work that applied smoothing to tag–anchor distances, we processed the absolute coordinate data directly.

### Data processing: preprocessing and missing data handling

With 27 tagged participants, all possible participant pairs (n = 351) were generated, and pairwise Euclidean distances were calculated at one-second intervals over the observation period. Because resident devices recorded positions less frequently than staff devices, resident coordinates were carried forward between observed time points to align pairwise resident-staff and resident-resident time series to a common one-second grid for operational comparison. Continuous missing periods of ≥3600 seconds were treated as invalid and excluded from analysis. This rule was selected because only a limited number of residents were independently mobile in this LTCF, and prolonged stationary periods may have reduced the availability of valid location data. Periods during which tags were clustered within a 50-centimetre radius in ≥10 participants (≥37% of all tagged participants) were excluded to account for temporary tag collection, such as charging or storage. These preprocessing steps were applied to all 27 participants.

### Contact classification

For each pair, distance at each second was binarised as 1 if the distance was ≤1.0 m and 0 otherwise. We then computed the total daily number of seconds meeting this distance threshold for each pair. Pairs with ≥900 seconds (15 minutes) of contact per day were classified as close contacts under the baseline operational definition. In sensitivity analyses, we evaluated alternative distance-time thresholds (1.0 m or 1.5 m combined with 5, 10, or 15 minutes) to assess how the operational definition affected directional discrepancies between questionnaire-derived records and UWB-derived proximity within this facility.

### Statistical analysis

Statistical analysis used UWB data from 16 residents and 10 staff members, and questionnaire data from the 10 staff who completed the survey. For each distance-time threshold, close contacts were identified separately from questionnaire-derived records and UWB-derived proximity. We quantified directional differences between the two sources using a discrepancy rate. For each study day, binary contact matrices were constructed separately for questionnaire-based records and UWB-derived proximity and compared within each contact pattern. Daily discrepancy was calculated at the participant level by summing row-wise differences between the two matrices and dividing by the number of eligible contacts for that participant within the relevant contact pattern on that day. Positive values indicated that questionnaire-based records identified fewer close contacts than UWB-derived proximity, whereas negative values indicated that questionnaire-based records identified more close contacts than UWB-derived proximity.

Analyses were stratified by contact pattern: (a) resident–resident contacts based on staff monitoring-based proxy records, (b) resident–staff contacts based on staff monitoring-based proxy records, (c) resident–staff contacts based on staff self-report, and (d) staff-staff contacts based on staff self-report. The pattern-specific units of analysis differed across contact patterns and are detailed in the Supplementary Methods and Table A3. Cross-pattern comparisons should therefore be interpreted descriptively as workflow-specific comparisons rather than as direct performance rankings across methods.

Because the study objective was operational comparison rather than population inference, analyses focused on the direction and magnitude of discrepancies between record-generation processes and a proximity-based operational measure within a single-facility workflow. In the staff self-report comparison, several alternative thresholds yielded discrepancies closer to zero than the baseline threshold in this dataset, but any apparent ranking across thresholds was treated as exploratory and metric-dependent rather than as identifying a generally applicable operational optimum. All statistical analyses were conducted in Python version 3.14.4 (Python Software Foundation).

## Results

Questionnaire-based records and UWB-derived proximity showed systematic but contact-type-specific discrepancies across the study period. As shown in Figure A1(Additional file 1), the histogram of interpersonal distance per second over the five-day survey showed a peak at around 1 to 2 meters for distances between residents and a peak at around 5 meters for distances between residents and facility staff. Distances in a range potentially relevant to operational close-contact assessment were therefore more commonly observed among resident-resident pairs than among resident-staff pairs in this dataset.

Figures 1A–1D compare the daily number of close contacts identified per participant by UWB-derived proximity and questionnaire-derived records, stratified by residents and staff. Full summary statistics are provided in Table A4 (Additional file 1). During the survey period, the number of contacts from questionnaires and UWB data was only slightly lower for residents, whereas greater discrepancies were observed among staff, with questionnaire-based counts tending to be higher than UWB-based counts.

**Figure 1.**
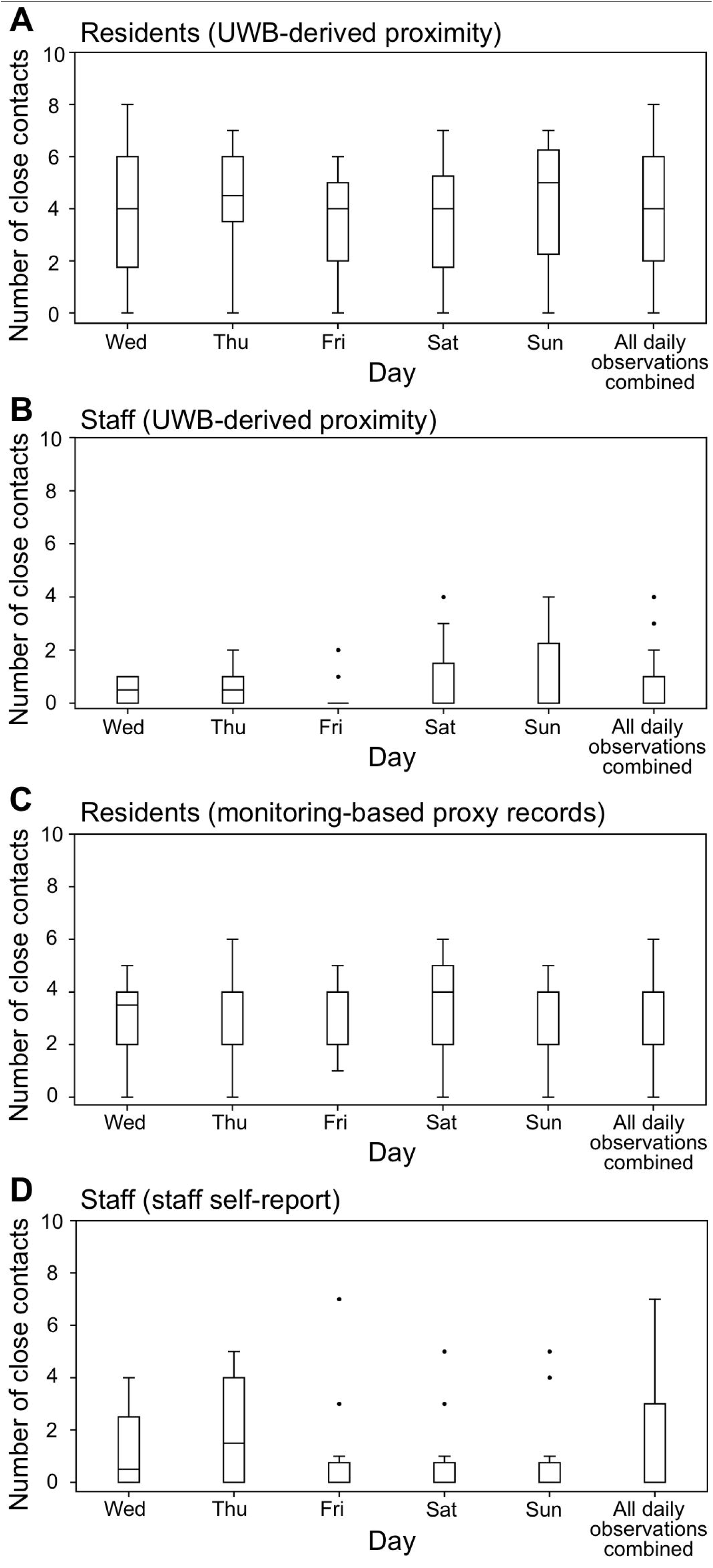
Daily number of close contacts (1.0 m, ≥15 min) per participant by role (UWB vs. Questionnaire) Close contacts were defined as individuals within 1.0 m for a cumulative total of ≥15 min during a day, regardless of mask use. Panels A–D compare the daily number of close contacts identified per participant by UWB-derived proximity and questionnaire-based records, stratified by residents and staff. Box plots show the median, interquartile range (IQR), and statistical outliers (black dots) for each study day (Wednesday to Sunday) and for all daily observations combined. Full summary statistics are provided in Table A4. Questionnaire-based resident contacts were derived from staff monitoring-based proxy records, whereas questionnaire-based staff contacts were derived from staff self-report.

Figures 2A–2C show the daily discrepancies between questionnaire-derived records and UWB-derived proximity by contact pattern under the baseline threshold of ≤1.0 m for ≥15 min [3–5]. Contact patterns are defined in the Methods and Supplementary Table A3. Pattern (d) was excluded because no close contacts were identified in either source during the study period. Patterns (b) and (c) were examined separately because they arose from different record-generation processes. Across the five study days, discrepancy patterns differed by contact type and by record-generation process: resident-resident contacts and resident-staff contacts based on monitoring-derived proxy records showed relatively small directional discrepancies, whereas resident-staff contacts based on staff self-report tended to show more negative discrepancies under the baseline threshold. These results should be interpreted as facility-specific comparisons of contact-list generation workflows rather than as validation against a transmission or diagnostic gold standard.

**Figure 2.**
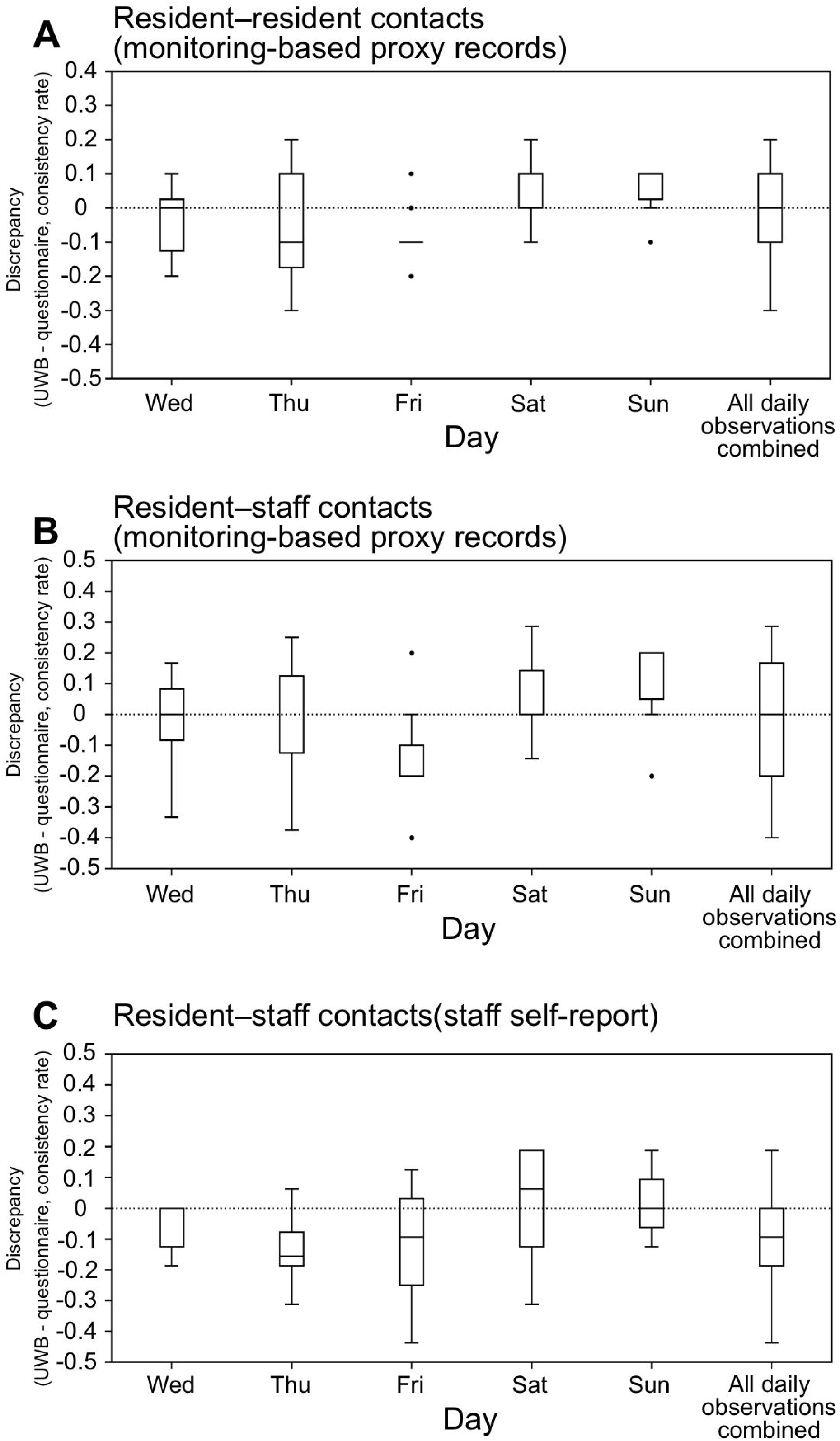
Daily discrepancies between UWB- and questionnaire-identified contacts (1.0 m, ≥15 min) by contact pattern: (A) resident–resident contacts based on staff monitoring-based proxy records, (B) resident–staff contacts based on staff monitoring-based proxy records, and (C) resident–staff contacts based on staff self-report. Box plots show the median, interquartile range (IQR), and statistical outliers (black dots) for each study day (Wednesday to Sunday) and for all daily observations combined. Positive values indicate that more contacts were detected by UWB-derived proximity than by questionnaire-based records, whereas negative values indicate that more contacts were identified by questionnaire-based records than by UWB-derived proximity. Panels represent (A) resident–resident contacts based on staff monitoring-based proxy records, (B) resident–staff contacts based on staff monitoring-based proxy records, and (C) resident–staff contacts based on staff self-report.

Sensitivity analyses showed that several alternative thresholds yielded discrepancies closer to zero than the baseline for staff self-reports of resident-staff contacts, although the apparent ranking varied by summary metric (Table A6; Figure 3). No single alternative threshold consistently minimised discrepancies across all summary metrics. This pattern may suggest that staff self-reported contacts captured shorter but operationally salient encounters not fully reflected by the baseline duration criterion. However, these findings should be interpreted as exploratory.

**Figure 3.**
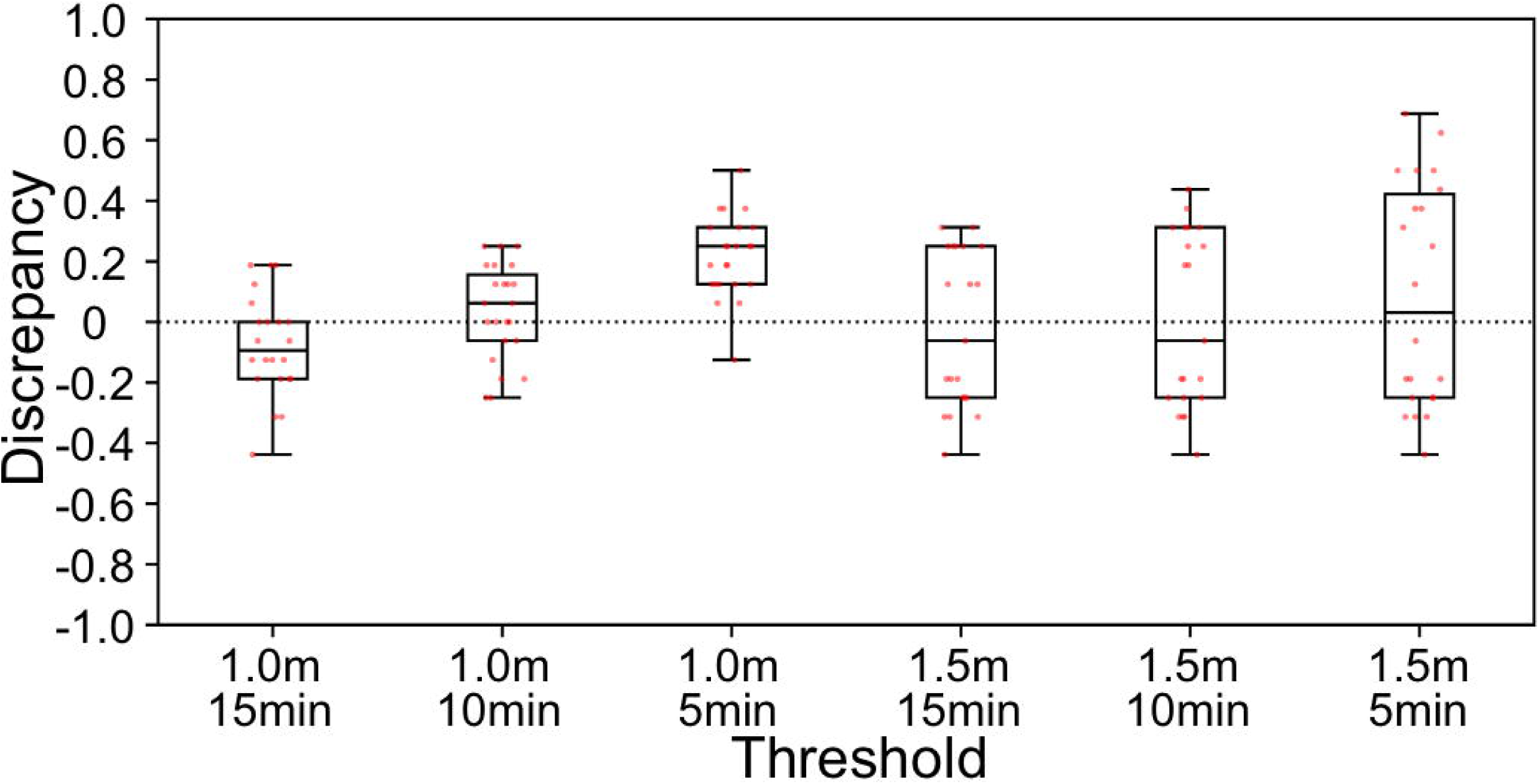
Sensitivity analysis of discrepancies under varying distance–time thresholds for self-reported contact with residents by staff. Each box plot shows the distribution of discrepancy values across all daily observations in the staff self-report comparison (contact pattern c) under each evaluated distance–time threshold. Box plots show the median, interquartile range (IQR), and statistical outliers, and red dots represent individual daily observations. Positive values indicate that more contacts were detected by UWB-derived proximity than by questionnaire-based records, whereas negative values indicate that more contacts were identified by questionnaire-based records than by UWB-derived proximity. Summary statistics for all thresholds are provided in Table A6

Supplementary Figure A2A–A2C show the cumulative proportion of unique close contacts identified by questionnaire-based records and UWB-derived proximity, counting backwards from the final survey day and stratified by contact pattern. In these analyses, cumulative percentages were normalised to the greater of the total number of unique contacts identified within 1.5 m for 10 minutes or more via UWB or questionnaire over the entire 5-day period, with this normalised to 100%. (See Supplementary Results.)

## Discussion

In this single-facility observational study in an LTCF, we compared two contact-list generation processes—staff self-report and monitoring-based proxy records—in relation to UWB-derived proximity. Questionnaire-derived records and UWB-derived proximity showed different patterns of alignment across contact types, indicating that the degree and direction of discrepancy depended on how contact lists were generated.

Our main finding was not simply that questionnaire-derived contacts differed from a proximity-based sensor measure, but that the discrepancy varied according to the record-generation process. Within this facility, resident-related monitoring-based proxy records showed relatively small directional discrepancies, whereas staff self-reported resident-staff contacts tended to identify additional contacts under the baseline threshold. The findings are therefore better interpreted as reflecting differences in workflow and documentation processes than as evidence that one approach is universally more accurate than another.

This distinction is especially relevant in LTCFs, where resident self-report is often limited or not feasible, and where contact lists may need to be assembled from staff observation, routine monitoring, and documentation embedded in care delivery. In such settings, monitoring-based proxy records are not merely substitutes for resident self-report; they are part of the operational workflow through which resident-related contacts are identified. Our results therefore support interpretation in terms of record-generation processes within LTCF practice rather than simple recall error alone.

By contrast, in our data, staff self-reports of resident-staff contacts tended to identify more contacts than were captured by the UWB-based operational definition under the baseline threshold. Similar discrepancies between self-reports and wearable sensor measurements have been reported in other settings [6, 7]. One plausible explanation is that staff may interpret “close contact” in operational or task-based terms, including direct-care episodes involving physical touch or face-to-face interaction, rather than as a strict cumulative distance-time measure, and may therefore report encounters that do not meet the questionnaire’s operational definition used in this study. In practice, this pattern could increase the number of contacts flagged for local review, which may affect testing, staffing, or cohorting decisions during outbreak response. Rather, apparent “overreporting” relative to a proximity-based threshold may partly reflect professional caution embedded in infection prevention and care practice, rather than memory error alone.

From an infection prevention and control perspective, this distinction may matter because more inclusive self-reported contact lists could increase the number of individuals considered for testing, work restriction, or cohorting, whereas monitoring-based records may provide a more feasible starting point for rapid local review in staffing-constrained LTCFs. However, practical implications should be interpreted cautiously because outbreak response effectiveness was not evaluated. Balancing completeness and feasibility of contact identification remains a key operational challenge in LTCFs.

Resident-resident contacts and resident-staff contacts based on monitoring-derived proxy records showed relatively small directional discrepancies in this dataset, whereas resident-staff contacts based on staff self-report more often exceeded the baseline proximity-based classification. Within this facility, these findings suggest that structured monitoring records may capture core resident-related contact patterns in a way that is broadly consistent with this proximity-based operational measure. This should not, however, be interpreted as validation against a true transmission standard. Rather, these findings suggest that monitoring-based records may serve as a stable and practical foundation for IPC-oriented contact identification in LTCFs.

In sensitivity analyses, several alternative thresholds yielded discrepancies closer to zero than the baseline, but no single threshold should be interpreted as operationally optimal because the apparent ranking varied by summary metric (Table A6). This pattern may indicate that staff self-reports captured shorter but operationally salient encounters in this workflow. However, these findings remain exploratory and should not be interpreted as identifying a generally applicable operational threshold. This reinforces the need for context-specific IPC definitions rather than universal thresholds.

With regard to implementation, our findings suggest a pragmatic hierarchy for contact identification in LTCFs. Monitoring-based records may serve as the primary operational tool for initial local review of resident-related contacts, while staff interviews remain an important complementary source, but may expand contact lists beyond those captured by the baseline proximity-based definition. This may be particularly relevant in staffing-constrained LTCFs, where rapid initial review of resident-related contacts is often required before a more detailed case-by-case assessment is possible. Where resources and local acceptability permit, sensor-informed review may be used as a supplementary facility-level reference for assessing how contact identification workflows function within a given LTCF. In this role, UWB-derived proximity should be interpreted as an operational measure of proximity rather than a direct measure of transmission risk, because it does not capture contextual modifiers such as ventilation, personal protective equipment use, orientation, or physical barriers. Implementation nevertheless requires attention to device management, privacy, trust, and feasibility, and strengthening existing monitoring workflows may still yield practical benefits.

This study has several limitations. First, the comparison relied on measurement conditions that were not fully symmetrical across participant groups. Staff and resident tags differed in temporal resolution because residents were monitored continuously over 24 hours, whereas overnight battery replacement was not possible because study personnel were not permitted to enter the facility between 20:00 and 07:00. Some resident tags were also mounted on wheelchairs or other supports, and resident coordinates were carried forward between observed time points to align pairwise time series to a one-second grid. In addition, prolonged missing periods and temporary tag clustering were excluded according to pre-specified preprocessing rules. These design features were operationally necessary in this facility but may have affected resident-related and staff-related classifications differently. Second, the study was conducted in a single facility over five days with a limited number of participants, and questionnaire data were available from 10 staff members only. The findings should therefore be interpreted as describing one facility-specific workflow rather than as estimating generalisable performance. Nevertheless, the high participation rate supports internal validity within this setting. Third, staff wore tags only while on duty, and both reporting behaviour and interpersonal behaviour may have been influenced by awareness of being monitored (a Hawthorne effect). Staff may have documented contacts more carefully than usual, and they may also have modified contact behaviour during routine care. Such effects may have been attenuated in longer observation periods, but they cannot be excluded in this five-day study. Fourth, UWB-derived proximity is not a measure of infection transmission and does not capture contextual factors that modify risk, such as ventilation, PPE use, body orientation, or physical barriers. Finally, the threshold sensitivity analysis used the same single-facility dataset and should therefore be regarded as exploratory and interpreted cautiously.

Future multi-site studies conducted over longer periods, and where feasible during actual outbreak investigations, are needed to determine whether similar workflow-specific patterns are observed across LTCFs and whether refined local operational definitions can support timely response while minimising avoidable isolation and workforce disruption. Future studies should also examine whether alignment between monitoring-based records, staff self-report, and UWB-derived proximity may vary according to residents’ care dependency, mobility, or ADL status, as such stratification could be relevant to practical infection prevention and control in LTCFs.

In conclusion, in this single-facility observational study, different contact-list generation processes were associated with different patterns of discrepancy relative to UWB-derived proximity. Within this facility, resident-related monitoring-based records showed relatively small directional discrepancies, whereas staff self-reports tended to identify additional resident-staff contacts under the baseline threshold. These findings suggest that, in LTCFs where resident self-report is often not feasible, monitoring-based records may support initial local contact-list review.

Sensor-informed review may also help facilities assess how their contact identification workflow relates to a proximity-based operational measure, but further evaluation in other LTCFs and in outbreak settings is needed. These findings should not be interpreted as establishing a generally applicable distance-time threshold beyond this facility. UWB-derived proximity may be most useful as a supplementary facility-level reference for reviewing contact identification workflows, rather than as a definitive measure of infection-relevant exposure.

## Data Availability

The individual-level UWB positioning data and questionnaire responses contain potentially identifiable information and will be used in ongoing graduate student research projects, including network analyses. For this reason, the full dataset cannot be shared publicly at this time. Deidentified summary statistics and analytic code supporting the findings of this article will be made available from the corresponding author upon publication. The full deidentified dataset will be deposited in a public repository within 2 years of publication. All authors had full access to the data and accept responsibility for the decision to submit for publication.

## Abbreviations

LTCF: Long-term care facilities
UWB: Ultra-wideband
IPC: Infection prevention and control
ADL: Activities of daily living

## CRediT authorship contribution statement

KM: Conceptualization, Data curation, Funding acquisition, Investigation, Methodology, Project administration, Resources, Writing – original draft. HS: Data curation, Formal analysis, Investigation, Methodology, Software, Validation, Visualization. Writing – original draft. All authors: Writing – review & editing. All authors approved the final version of the manuscript.

## Ethics statement

The study protocol was reviewed and approved by the Ethics Committee of Kyoto University Graduate School of Medicine (Approval No. R6-G-8-2). Written informed consent was obtained from participating staff for both the questionnaire and location measurements. For residents who only contributed location measurements, informed consent was obtained via an opt-out procedure after prior notification.

## Data Availability Statement

The datasets generated and/or analysed during the current study are not publicly available because they contain potentially identifiable location and contact data from residents and staff in a single long-term care facility. De-identified data may be made available from the corresponding author on reasonable request, subject to review and approval by the relevant ethics committee and data-sharing agreements in accordance with institutional and legal requirements.

## Acknowledgments

The authors thank the participating long-term care facility, its administrative leadership, and staff members for their cooperation and support in conducting this study.

## Declaration of generative AI and AI-assisted technologies in the writing process

During the preparation of this manuscript, the authors used ChatGPT (OpenAI) to improve the clarity and readability of the text. All AI-assisted output was reviewed and edited by the authors, who take full responsibility for the final content of the manuscript.

## Supplementary Information

The following supplementary materials are provided with this article: Additional file 1. Supplementary Methods, Tables A1–A6, and Figures A1–A2. Additional file 2. Questionnaire survey (English translation).

## References

1. European Centre for Disease Prevention and Control (ECDC). Surveillance of COVID-19 in long-term care facilities in the EU/EEA. Stockholm: ECDC; 2021. Available from: https://www.ecdc.europa.eu/en/publications-data/surveillance-covid-19-long-term-care-facilities-eueea (Accessed 2025 Dec 21).

2. Centers for Disease Control and Prevention (CDC). Investigating a COVID-19 case: interim guidance on developing a COVID-19 case investigation & contact tracing plan. Updated 21 Jan 2022. Available from: https://archive.cdc.gov/www_cdc_gov/coronavirus/2019-ncov/php/contact-tracing/contact-tracing-plan/investigating-covid-19-case.html (Accessed 2025 Dec 21).

3. National Institute of Infectious Diseases (Japan). Provisional guidelines for active epidemiological investigation for COVID-19 (17 Jan 2020 and 21 Jan 2020 versions). Available from: https://id-info.jihs.go.jp (Accessed 2025 Dec 21).

4. National Institute of Infectious Diseases (Japan). Provisional guidelines for active epidemiological investigation for COVID-19 (20 Apr 2020 version). Available from: https://id-info.jihs.go.jp (Accessed 2025 Dec 21).

5. National Institute of Infectious Diseases (Japan). Q&A on the revision of the definition of close contacts in the provisional guidelines for active epidemiological investigation (27 Apr 2020). Available from: https://id-info.jihs.go.jp (Accessed 2025 Dec 21).

6. Mastrandrea R, Fournet J, Barrat A. Contact patterns in a high school: a comparison between data collected using wearable sensors, contact diaries and friendship surveys. PLoS One. 2015;10(9):e0136497. doi:10.1371/journal.pone.0136497.

7. Smieszek T, Castell S, Barrat A, Cattuto C, White PJ, Krause G. Contact diaries versus wearable proximity sensors in measuring contact patterns at a conference: method comparison and participants’ attitudes. BMC Infect Dis. 2016;16:341. doi:10.1186/s12879-016-1676-y.

8. Qian L, Chan A, Cai J, Lewicke J, Gregson G, Lipsett M, et al. Evaluation of the accuracy of a UWB tracker for in-home positioning for older adults. Med Eng Phys. 2024;126:104155. doi:10.1016/j.medengphy.2024.104155.

9. Alarifi A, Al-Salman A, Alsaleh M, Alnafessah A, Al-Hadhrami S, Al-Ammar MA, et al. Ultra wideband indoor positioning technologies: analysis and recent advances. Sensors (Basel). 2016;16(5):707. doi:10.3390/s16050707.

10. Shinto H, Ohki Y, Mizumoto K, Saito K. Visualization of interpersonal communication using indoor positioning technology with UWB tags. arXiv. 2025. doi:10.48550/arXiv.2510.06797.

11. Duval A, Obadia T, Martinet L, Boëlle PY, Fleury E, Guillemot D, et al. Measuring dynamic social contacts in a rehabilitation hospital: effect of wards, patient and staff characteristics. Sci Rep. 2018;8:1686. doi:10.1038/s41598-018-20008-w.

12. Stokes K, Piaggio D, De Micco F, Zarro M, De Benedictis A, Tambone V, et al. The use of contact tracing technologies for infection prevention and control purposes in nosocomial settings: a systematic literature review. Infect Dis Rep. 2024;16:519–530. doi:10.3390/idr16030039.

13. Thompson CA, Willis TA, Farrin A, Gordon A, Daffu-O’Reilly A, Noakes C, et al. Technology-enabled CONTACT tracing in care homes in the COVID-19 pandemic: the CONTACT non-randomised mixed-methods feasibility study. Health Technol Assess. 2025;29(24):1–24. doi:10.3310/UHDN6497.

