## Supplementary information 1 for "Monitoring-based and self-reported close-contact records in relation to ultra-wideband-derived proximity in a long-term care facility: a single-facility observational study"

The electronic supplementary material is below the table titles.

Supplementary Methods, Results, Tables A1–A6, and Figure A1–A2 provide supporting data referenced in the main text.

### Supplementary Methods

Participants wore UWB tags, but sampling frequency differed by participant group because of operational constraints in the facility. Staff tags were recorded at 1 Hz (once every second), whereas resident tags were recorded once every 10 seconds because residents wore tags continuously over 24 hours and study personnel were not permitted to enter the facility between 20:00 and 07:00, precluding overnight battery replacement. Interpersonal distances were analysed on a one-second grid based on the estimated (x, y) positions of each tag. For resident-related pairs, the most recent observed resident coordinate was carried forward between recorded time points to align pairwise time series for operational comparison. Periods with missing or implausible coordinates were excluded according to pre-specified rules (Table A2).

Definition of close contact: Close contact events were defined using the official distance–time threshold (within 1.0 m for ≥15 minutes) as the primary definition. Sensitivity analyses evaluated alternative thresholds (1.0 m/1.5 m × 5/10/15 min) (Table A6).

Construction of contact matrices (UWB): For each day and each contact pattern, we constructed a binary adjacency matrix A_UWB where element (i, j)=1 indicates that pair (i, j) met the close contact definition on that day according to UWB measurements, otherwise 0. UWB-based contacts were treated as bidirectional.

Construction of contact matrices (staff report): Staff-reported contacts were extracted from (i) routine monitoring history logs (proxy reports) and (ii) staff self-reported questionnaires, depending on the contact pattern (Table A3). When reports were directional, matrices were symmetrised for pairwise comparisons (i.e. if either member of a pair was reported as a close contact, both corresponding cells were set to 1).

Consistency rate (discrepancy) between methods: For each day, we computed a discrepancy matrix D = A_UWB − A_report. Each element takes values +1 (contact detected only by UWB; underreporting), −1 (only by report; overreporting), or 0 (detected by both). For each participant i, we calculated the row-wise mean discrepancy over all eligible counterparts, yielding a daily consistency rate. Positive values indicate net underreporting relative to UWB; negative values indicate net overreporting.

Analysis unit: Discrepancies were summarised by day and by contact pattern (Table A5) and used for threshold sensitivity analyses (Table A6).

### Supplementary Results

Supplementary Figure A2A–A2C illustrate how the cumulative proportion of unique close contacts changed when questionnaire-based records and UWB-derived proximity were counted backward from the final survey day, stratified by contact pattern. The cumulative pattern varied by contact type, and resident–resident contacts based on staff monitoring-based proxy records levelled off relatively early within the observation period. In all panels, the cumulative percentage is normalized to the greater of the total number of unique contacts identified at a distance of 1.5m or less for 10 minutes or more over the entire 5-day period by UWB or questionnaire (set to 100%).

Supplementary Figure A1 showed that distances potentially relevant to operational close-contact assessment were more commonly observed among resident–resident pairs than among resident–staff pairs under observed conditions. Figures A2A and A2B show discrepancies close to zero, but the cumulative results indicate substantial differences between UWB-derived and questionnaire-based records in the number of unique contact pairs identified over time. Of particular importance, the total number of unique close-contact combinations identified by questionnaire-based records was lower than that identified by UWB. Because the discrepancy summaries do not address unique combinations, the two sources might appear broadly consistent at first glance. However, UWB identified a greater number of unique combinations than questionnaire-based records in these panels. This pattern may indicate that UWB captured additional combinations that were not represented in questionnaire-based records and that integration of multiple staff monitoring-based proxy records may still incompletely capture all unique pairings. Specifically, in Figure A2A, resident–resident contacts identified from staff monitoring-based proxy records levelled off by day 2 and showed little change thereafter, whereas contacts detected by UWB continued to increase until day 5. One possible explanation is that staff monitoring-based proxy records more closely reflected relatively stable resident placement and observation patterns than the time-varying proximity measured by UWB. In Figure A2B, resident–staff contacts measured by UWB increased rapidly in the most recent 1–2 days, whereas contacts based on staff monitoring-based proxy records increased more gradually, only roughly matching the UWB gradient in the latter half of the 5-day period. This pattern may suggest that staff monitoring-based proxy records reflected routine care or observation patterns aggregated across days more strongly than time-specific proximity episodes captured by UWB under the baseline threshold.

In Figure A2C, staff self-reported resident–staff contacts were similar to UWB-derived contacts over the most recent one to two days, but continued to accumulate thereafter and exceeded UWB-derived contacts over longer recall windows, with the gap widening toward day 5. One possible explanation is that staff may have reported operationally salient resident contacts, including repeated brief care episodes or face-to-face interactions, even when cumulative time within 1 m did not meet the baseline duration criterion used here. This interpretation is consistent with the observation that several alternative thresholds were associated with discrepancies closer to zero than the baseline in this dataset, although the apparent ranking varied by summary metric, and should therefore be regarded as exploratory and facility-specific.

### Supplementary Tables A1–A6

**Table A1. Participant characteristics and questionnaire/monitoring coverage.**

| Role | Eligible (n) | Consented & wore UWB tags (n) | Included in UWB analysis (n) | Staff questionnaire available (n) | Monitoring report available (n) |
| --- | --- | --- | --- | --- | --- |
| Residents | 18 | 16 | 16 | N/A | 16 |
| Facility staff | 11 | 11 | 11 | 10 | 10  (for residents) |
| Total | 29 | 27 | 27 | 10 | 16 residents |
| Notes: Staff wore UWB tags during working hours; residents wore tags continuously during the survey period. Questionnaire-based self-report was collected from staff only. | | | | | |

**Table A2. UWB system configuration and data processing parameters.**

| Parameter | Value | Notes |
| --- | --- | --- |
| Sampling frequency (staff tags) | 1 Hz (once every second) | UWB tags transmitted at one-second intervals. |
| Sampling frequency (resident tags) | 0.1 Hz (once every 10 seconds) | Resident tags were recorded less frequently to permit continuous 24-hour wear during the study period. Overnight battery replacement was not possible because study personnel were not permitted to enter the facility between 20:00 and 07:00. |
| Number of UWB receivers | 42 | Receivers were installed throughout the facility. |
| Location smoothing | Kalman filter + LSTM (120-s window) | Used to reduce noise in raw coordinates. |
| Invalid periods (missing data) | Excluded if missing ≥3600 s^§^ | Periods without valid coordinates were removed. |
| Crowding exclusion | Excluded when ≥10 participants within 50 cm | To mitigate artefacts from tag collision/multipath. |
| Primary close contact definition | ≤1.0 m for ≥15 min | Official threshold used for main analyses. |
| Sensitivity thresholds | 1.0 m/1.5 m × 5/10/15 min | Evaluated in sensitivity analyses (Table A6). |
| Note: The 3600-s threshold was selected because only a limited number of residents were independently mobile in this LTCF, and prolonged lack of movement may have reduced continued reception of tag transmissions by UWB receivers, resulting in missing location data.  Because sampling frequency and tag placement differed between staff and residents, UWB-derived proximity should be interpreted as a facility-specific operational measure in this study. | | |

**Table A3. Definition of contact patterns and data sources used for comparison.**

| Contact pattern | Pair type | Report type | Reported by | Compared with | Unit of analysis |
| --- | --- | --- | --- | --- | --- |
| a | Resident–Resident | Proxy report (monitoring history) | Staff | UWB proximity between residents | Residents (per day) |
| b | Residents–Staff | Proxy report (monitoring history) | Staff | UWB proximity between residents and staff | Residents / staff (per day) |
| c | Residents–Staff | Self-report questionnaire | Staff | UWB proximity between residents and staff | Staff member (per day) |
| d | Staff –Staff | Self-report questionnaire | Staff | UWB proximity between staff | Not analysed |
| Notes: Contact pattern d (staff–staff) was specified a priori but was not analysed further because no close contacts were identified for this pattern in the comparative analyses. | | | | | |

**Table A4. Daily number of close contacts per participant by role and method (UWB vs questionnaire) corresponding to Figure 1A –1D.**

| Role | Method | Number of close contacts  (Mean, median, IQR) | | | | | |
| --- | --- | --- | --- | --- | --- | --- | --- |
|  |  | Wednesday | Thursday | Friday | Saturday | Sunday | All daily observations combined |
| Resident | UWB | (3.9, 4.0, 1.8–6.0) | (4.1, 4.5, 3.5–6.0) | (3.4, 4.0, 2.0–5.0) | (3.6, 4.0, 1.8–5.3) | (4.0, 5.0, 2.3–6.3) | (3.8, 4.0, 2.0–6.0) |
| Resident | Questionnaire (proxy report) | (3.1, 3.5, 2.0–4.0) | (3.4, 4.0, 2.0–4.0) | (3.4, 4.0, 2.0–4.0) | (3.4, 4.0, 2.0–5.0) | (3.2, 4.0, 2.0–4.0) | (3.3, 4.0, 2.0-4.0) |
| Staff | UWB | (0.5, 0.5, 0.0–1.0) | (0.7, 0.5, 0.0–1.0) | (0.3, 0.0, 0.0–0.0) | (0.9, 0.0, 0.0–1.5) | (1.0, 0.0, 0.0–2.3) | (0.68, 0.0, 0.0-1.0) |
| Staff | Questionnaire (self-report) | (1.2, 0.5, 0.0–2.5) | (2.0, 1.5, 0.0–4.0) | (1.1, 0.0, 0.0–0.75) | (0.9, 0.0, 0.0–0.75) | (1.0, 0.0, 0.0–0.75) | (1.2, 0.0, 0.0-3.0) |

**Table A5. Daily discrepancies between UWB- and questionnaire-identified close contacts under the baseline threshold (1.0 m, ≥15 min) by contact pattern, corresponding to Figure 2A–2C.**

| Contact pattern | Statistic | Wednesday | Thursday | Friday | Saturday | Sunday | All daily observations combined |
| --- | --- | --- | --- | --- | --- | --- | --- |
| a | Mean | 0.062 | 0.058 | 0.036 | -0.0044 | 0.036 | 0.037 |
| a | Median | 0.0 | 0.0 | 0.0 | 0.0 | 0.0 | 0.0 |
| a | IQR | -0.033 to 0.17 | 0.0 to 0.1 | -0.033 to 0.1 | -0.1 to 0.1 | -0.067 to 0.17 | -0.067 to 0.13 |
| b | Mean | -0.024 | -0.042 | -0.13 | 0.048 | 0.1 | -0.011 |
| b | Median | 0.0 | -0.13 | -0.2 | 0.0 | 0.2 | 0.0 |
| b | IQR | -0.083 to 0.083 | -0.13 to 0.13 | -0.2 to -0.1 | 0.0 to 0.14 | 0.05 to 0.2 | -0.2 to 0.17 |
| c | Mean | -0.088 | -0.14 | -0.13 | 0.0 | 0.021 | -0.077 |
| c | Median | -0.13 | -0.16 | -0.094 | 0.063 | 0.0 | -0.094 |
| c | IQR | -0.13 to 0.0 | -0.19 to -0.078 | -0.25 to -0.031 | -0.13 to 0.19 | -0.063 to 0.094 | -0.19 to 0.0 |
| Notes: Results are shown separately for contact patterns a–c, corresponding to Figure 2A–2C in the main manuscript. Discrepancy was defined as UWB-based classification minus questionnaire-based classification. Positive values indicate underreporting in questionnaire-based records relative to UWB-derived proximity, whereas negative values indicate overreporting. IQRs are presented from the lower to the upper quartile. | | | | | | | |

**Table A6. Discrepancies under alternative distance–time thresholds for the staff self-report comparison (contact pattern c), corresponding to Figure 3.**

| Threshold | Mean discrepancy | Median discrepancy | IQR |
| --- | --- | --- | --- |
| 1.0 m, ≥15 min (primary) | -0.077 | -0.094 | -0.19 to 0.0 |
| 1.0 m, ≥10 min | 0.035 | 0.063 | -0.063 to 0.16 |
| 1.0 m, ≥5 min | 0.22 | 0.25 | 0.13 to 0.31 |
| 1.5 m, ≥15 min | -0.024 | -0.063 | -0.25 to 0.25 |
| 1.5 m, ≥10 min | 0.0089 | -0.063 | -0.25 to 0.31 |
| 1.5 m, ≥5 min | 0.088 | 0.031 | -0.25 to 0.42 |
| Notes: Results are shown for the staff self-report comparison (contact pattern c). Discrepancy was defined as UWB-based classification minus questionnaire-based classification. Positive values indicate underreporting in questionnaire-based records relative to UWB-derived proximity, whereas negative values indicate overreporting. IQRs are presented from the lower to the upper quartile.  Several alternative thresholds were associated with discrepancies closer to zero than the baseline in this dataset, but the apparent ranking varied by summary metric; these findings should therefore be interpreted as exploratory and facility-specific rather than as identifying a single operationally optimal threshold. | | | |

**Supplementary Figures A1–A2**

**Figure A1. Distribution of one-second pairwise distances across the full survey period, stratified by participant-pair type (resident-resident, resident-staff, and staff-staff).**

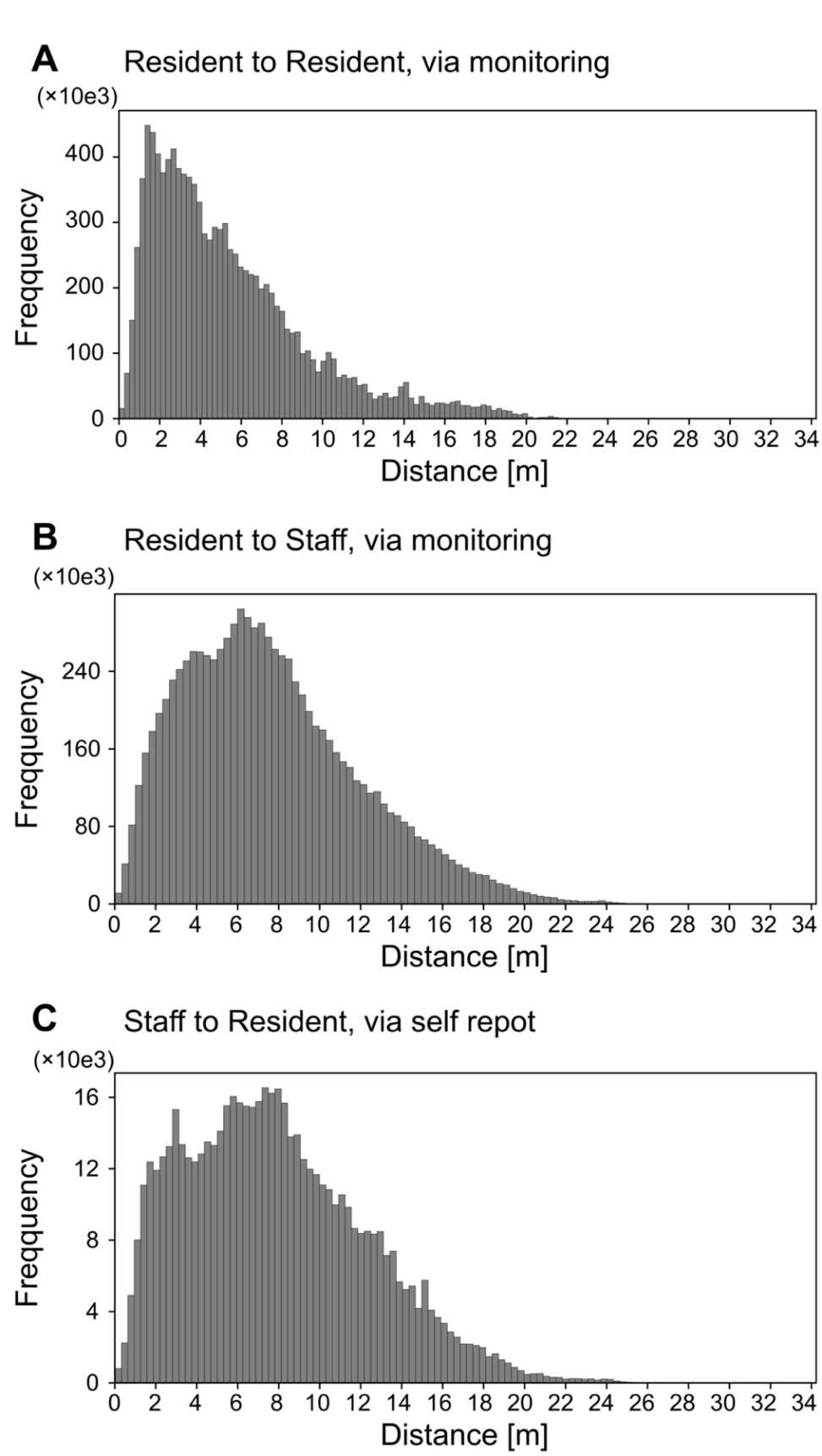

**Figure A2. Cumulative proportion of unique close contacts identified by questionnaire-based records and UWB-derived proximity, counting backward from the final survey day and stratified by contact pattern: (A) resident–resident contacts based on staff monitoring-based proxy records, (B) resident–staff contacts based on staff monitoring-based proxy records, and (C) resident–staff contacts based on staff self-report.**

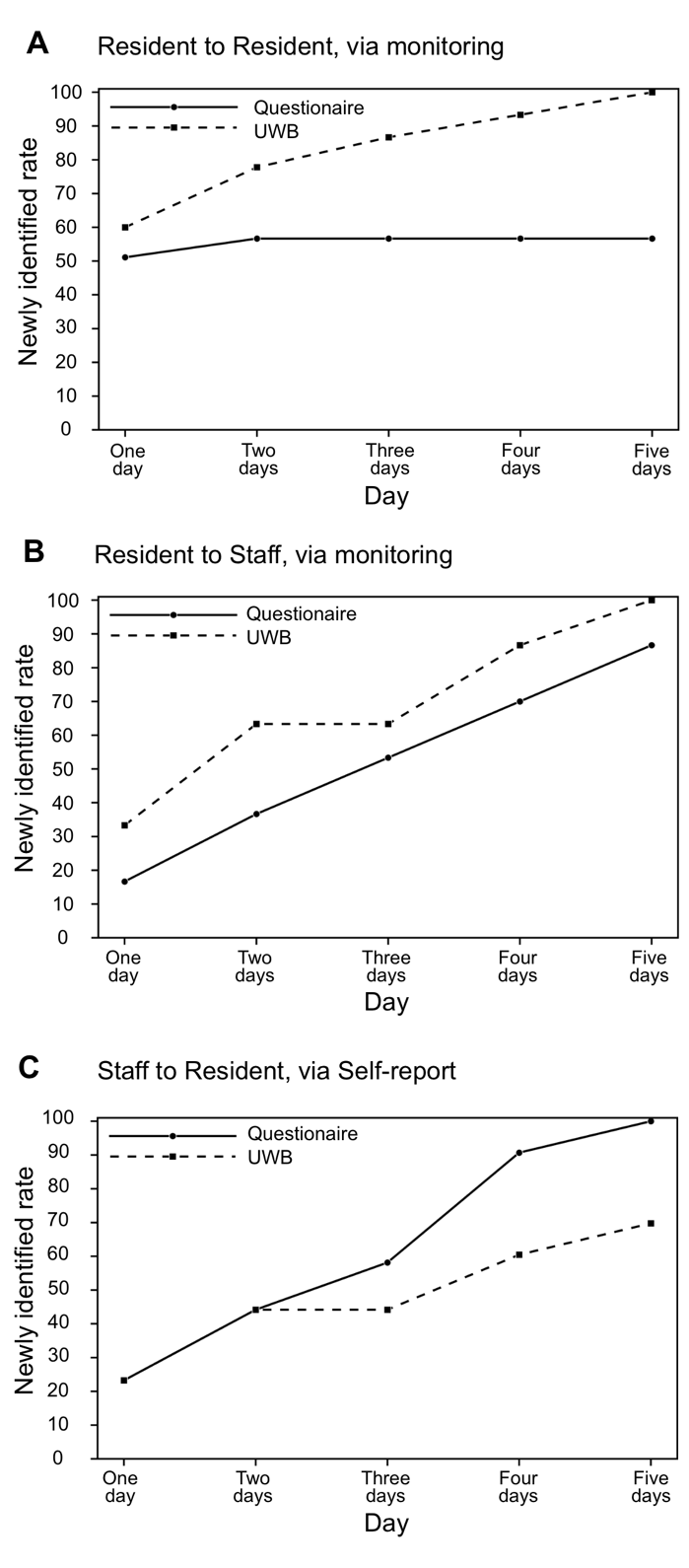

In all panels, the cumulative percentage is normalized to the greater of the total number of unique contacts identified by UWB or questionnaires for 10 minutes or more within 1.5 m over the entire 5-day period (set to 100%).
