## Supplementary information 2 for "Monitoring-based and self-reported close-contact records in relation to ultra-wideband-derived proximity in a long-term care facility: a single-facility observational study"

**Long-term care facility questionnaire (staff version)**

**English Translation**

**Date completed: December ___, 2024**

**Q1. (Refer to the ID reference table) Please provide your ID number.**

**(	)**

**Q2. Please indicate your gender by circling the applicable option.**

**Male / Female / Other**

**Q3. Please indicate your age.**

**( ) years**

**Q4. Please indicate your years of service at this facility.**

**( ) years**

**Q5. Within the designated area and during the designated period, please circle the ID numbers of individuals who qualify as your “close contacts.” Responses are organised by date. The definitions of “close contact,” “designated area,” and “designated period” are as follows.**

- **“Close contact”: An individual with whom you had contact within 1 metre for a cumulative total of 15 minutes or longer during the designated period, regardless of mask use.**
- **“Designated period”: The period during which you wore the tag (i.e. your working hours within the study period from 9:00 AM on Wednesday, December 11, 2024, to 9:00 AM on Monday, December 16, 2024).**

**“Designated area”: The Mitsuba and Basil units, including restrooms and bath areas.**

| **ID** | **Role** | **Unite** | **Wed, Dec 11**  **After 9:00 AM** | **Thu,Dec 12** | **Fri,Dec 13** | **Sat,Dec 14** | **Sun, Dec 15** | **Mon, Dec 16**  **Before 9:00AM** |
| --- | --- | --- | --- | --- | --- | --- | --- | --- |
| 1 | **Resident** | Mitsuba |  |  |  |  |  |  |
| 2 | **Resident** | Mitsuba |  |  |  |  |  |  |
| 3 | **Resident** | Mitsuba |  |  |  |  |  |  |
| 4 | **Resident** | Mitsuba |  |  |  |  |  |  |
| 5 | **Resident** | Mitsuba |  |  |  |  |  |  |
| 6 | **Resident** | Mitsuba |  |  |  |  |  |  |
| 7 | **Resident** | Mitsuba |  |  |  |  |  |  |
| 8 | **Resident** | Mitsuba |  |  |  |  |  |  |
| 9 | **Resident** | Mitsuba |  |  |  |  |  |  |
| 10 | **Resident** | **Basil** |  |  |  |  |  |  |
| 11 | **Resident** | **Basil** |  |  |  |  |  |  |
| 12 | **Resident** | **Basil** |  |  |  |  |  |  |
| 13 | **Resident** | **Basil** |  |  |  |  |  |  |
| 14 | **Resident** | **Basil** |  |  |  |  |  |  |
| 15 | **Resident** | **Basil** |  |  |  |  |  |  |
| 16 | **Resident** | **Basil** |  |  |  |  |  |  |
| 17 | **Resident** | **Basil** |  |  |  |  |  |  |
| 18 | **Resident** | **Basil** |  |  |  |  |  |  |
| 19 | Staff | Mitsuba |  |  |  |  |  |  |
| 20 | Staff | Mitsuba |  |  |  |  |  |  |
| 21 | Staff | Mitsuba |  |  |  |  |  |  |
| 22 | Staff | Mitsuba |  |  |  |  |  |  |
| 23 | Staff | Mitsuba |  |  |  |  |  |  |
| 24 | Staff | Mitsuba |  |  |  |  |  |  |
| 25 | Staff | **Basil** |  |  |  |  |  |  |
| 26 | Staff | **Basil** |  |  |  |  |  |  |
| 27 | Staff | **Basil** |  |  |  |  |  |  |
| 28 | Staff | **Basil** |  |  |  |  |  |  |
| 29 | Staff | **Basil** |  |  |  |  |  |  |

**Q6. Within the designated area and during the designated period, please circle the ID numbers of any individuals who qualify as close contacts of the following resident. Responses are organised by date. The definitions of “close contact,” “designated area,” and “designated period” are the same as above.**

**Resident ID	1**

| **ID** | **Role** | **Unite** | **Wed, Dec 11**  **After 9:00 AM** | **Thu,Dec 12** | **Fri,Dec 13** | **Sat,Dec 14** | **Sun, Dec 15** | **Mon, Dec 16**  **Before 9:00AM** |
| --- | --- | --- | --- | --- | --- | --- | --- | --- |
| 1 | **Resident** | Mitsuba |  |  |  |  |  |  |
| 2 | **Resident** | Mitsuba |  |  |  |  |  |  |
| 3 | **Resident** | Mitsuba |  |  |  |  |  |  |
| 4 | **Resident** | Mitsuba |  |  |  |  |  |  |
| 5 | **Resident** | Mitsuba |  |  |  |  |  |  |
| 6 | **Resident** | Mitsuba |  |  |  |  |  |  |
| 7 | **Resident** | Mitsuba |  |  |  |  |  |  |
| 8 | **Resident** | Mitsuba |  |  |  |  |  |  |
| 9 | **Resident** | Mitsuba |  |  |  |  |  |  |
| 10 | **Resident** | **Basil** |  |  |  |  |  |  |
| 11 | **Resident** | **Basil** |  |  |  |  |  |  |
| 12 | **Resident** | **Basil** |  |  |  |  |  |  |
| 13 | **Resident** | **Basil** |  |  |  |  |  |  |
| 14 | **Resident** | **Basil** |  |  |  |  |  |  |
| 15 | **Resident** | **Basil** |  |  |  |  |  |  |
| 16 | **Resident** | **Basil** |  |  |  |  |  |  |
| 17 | **Resident** | **Basil** |  |  |  |  |  |  |
| 18 | **Resident** | **Basil** |  |  |  |  |  |  |
| 19 | Staff | Mitsuba |  |  |  |  |  |  |
| 20 | Staff | Mitsuba |  |  |  |  |  |  |
| 21 | Staff | Mitsuba |  |  |  |  |  |  |
| 22 | Staff | Mitsuba |  |  |  |  |  |  |
| 23 | Staff | Mitsuba |  |  |  |  |  |  |
| 24 | Staff | Mitsuba |  |  |  |  |  |  |
| 25 | Staff | **Basil** |  |  |  |  |  |  |
| 26 | Staff | **Basil** |  |  |  |  |  |  |
| 27 | Staff | **Basil** |  |  |  |  |  |  |
| 28 | Staff | **Basil** |  |  |  |  |  |  |
| 29 | Staff | **Basil** |  |  |  |  |  |  |

**Q7. Within the designated area and during the designated period, please circle the ID numbers of any individuals who qualify as close contacts of the following resident. Responses are organised by date. The definitions of “close contact,” “designated area,” and “designated period” are the same as above.**

**Resident ID	2**

| **ID** | **Role** | **Unite** | **Wed, Dec 11**  **After 9:00 AM** | **Thu,Dec 12** | **Fri,Dec 13** | **Sat,Dec 14** | **Sun, Dec 15** | **Mon, Dec 16**  **Before 9:00AM** |
| --- | --- | --- | --- | --- | --- | --- | --- | --- |
| 1 | **Resident** | Mitsuba |  |  |  |  |  |  |
| 2 | **Resident** | Mitsuba |  |  |  |  |  |  |
| 3 | **Resident** | Mitsuba |  |  |  |  |  |  |
| 4 | **Resident** | Mitsuba |  |  |  |  |  |  |
| 5 | **Resident** | Mitsuba |  |  |  |  |  |  |
| 6 | **Resident** | Mitsuba |  |  |  |  |  |  |
| 7 | **Resident** | Mitsuba |  |  |  |  |  |  |
| 8 | **Resident** | Mitsuba |  |  |  |  |  |  |
| 9 | **Resident** | Mitsuba |  |  |  |  |  |  |
| 10 | **Resident** | **Basil** |  |  |  |  |  |  |
| 11 | **Resident** | **Basil** |  |  |  |  |  |  |
| 12 | **Resident** | **Basil** |  |  |  |  |  |  |
| 13 | **Resident** | **Basil** |  |  |  |  |  |  |
| 14 | **Resident** | **Basil** |  |  |  |  |  |  |
| 15 | **Resident** | **Basil** |  |  |  |  |  |  |
| 16 | **Resident** | **Basil** |  |  |  |  |  |  |
| 17 | **Resident** | **Basil** |  |  |  |  |  |  |
| 18 | **Resident** | **Basil** |  |  |  |  |  |  |
| 19 | Staff | Mitsuba |  |  |  |  |  |  |
| 20 | Staff | Mitsuba |  |  |  |  |  |  |
| 21 | Staff | Mitsuba |  |  |  |  |  |  |
| 22 | Staff | Mitsuba |  |  |  |  |  |  |
| 23 | Staff | Mitsuba |  |  |  |  |  |  |
| 24 | Staff | Mitsuba |  |  |  |  |  |  |
| 25 | Staff | **Basil** |  |  |  |  |  |  |
| 26 | Staff | **Basil** |  |  |  |  |  |  |
| 27 | Staff | **Basil** |  |  |  |  |  |  |
| 28 | Staff | **Basil** |  |  |  |  |  |  |
| 29 | Staff | **Basil** |  |  |  |  |  |  |

**Q8. Within the designated area and during the designated period, please circle the ID numbers of any individuals who qualify as close contacts of the following resident. Responses are organised by date. The definitions of “close contact,” “designated area,” and “designated period” are the same as above.**

**Resident ID	3**

| **ID** | **Role** | **Unite** | **Wed, Dec 11**  **After 9:00 AM** | **Thu,Dec 12** | **Fri,Dec 13** | **Sat,Dec 14** | **Sun, Dec 15** | **Mon, Dec 16**  **Before 9:00AM** |
| --- | --- | --- | --- | --- | --- | --- | --- | --- |
| 1 | **Resident** | Mitsuba |  |  |  |  |  |  |
| 2 | **Resident** | Mitsuba |  |  |  |  |  |  |
| 3 | **Resident** | Mitsuba |  |  |  |  |  |  |
| 4 | **Resident** | Mitsuba |  |  |  |  |  |  |
| 5 | **Resident** | Mitsuba |  |  |  |  |  |  |
| 6 | **Resident** | Mitsuba |  |  |  |  |  |  |
| 7 | **Resident** | Mitsuba |  |  |  |  |  |  |
| 8 | **Resident** | Mitsuba |  |  |  |  |  |  |
| 9 | **Resident** | Mitsuba |  |  |  |  |  |  |
| 10 | **Resident** | **Basil** |  |  |  |  |  |  |
| 11 | **Resident** | **Basil** |  |  |  |  |  |  |
| 12 | **Resident** | **Basil** |  |  |  |  |  |  |
| 13 | **Resident** | **Basil** |  |  |  |  |  |  |
| 14 | **Resident** | **Basil** |  |  |  |  |  |  |
| 15 | **Resident** | **Basil** |  |  |  |  |  |  |
| 16 | **Resident** | **Basil** |  |  |  |  |  |  |
| 17 | **Resident** | **Basil** |  |  |  |  |  |  |
| 18 | **Resident** | **Basil** |  |  |  |  |  |  |
| 19 | Staff | Mitsuba |  |  |  |  |  |  |
| 20 | Staff | Mitsuba |  |  |  |  |  |  |
| 21 | Staff | Mitsuba |  |  |  |  |  |  |
| 22 | Staff | Mitsuba |  |  |  |  |  |  |
| 23 | Staff | Mitsuba |  |  |  |  |  |  |
| 24 | Staff | Mitsuba |  |  |  |  |  |  |
| 25 | Staff | **Basil** |  |  |  |  |  |  |
| 26 | Staff | **Basil** |  |  |  |  |  |  |
| 27 | Staff | **Basil** |  |  |  |  |  |  |
| 28 | Staff | **Basil** |  |  |  |  |  |  |
| 29 | Staff | **Basil** |  |  |  |  |  |  |

**Q9. Within the designated area and during the designated period, please circle the ID numbers of any individuals who qualify as close contacts of the following resident. Responses are organised by date. The definitions of “close contact,” “designated area,” and “designated period” are the same as above.**

**Resident ID	4**

| **ID** | **Role** | **Unite** | **Wed, Dec 11**  **After 9:00 AM** | **Thu,Dec 12** | **Fri,Dec 13** | **Sat,Dec 14** | **Sun, Dec 15** | **Mon, Dec 16**  **Before 9:00AM** |
| --- | --- | --- | --- | --- | --- | --- | --- | --- |
| 1 | **Resident** | Mitsuba |  |  |  |  |  |  |
| 2 | **Resident** | Mitsuba |  |  |  |  |  |  |
| 3 | **Resident** | Mitsuba |  |  |  |  |  |  |
| 4 | **Resident** | Mitsuba |  |  |  |  |  |  |
| 5 | **Resident** | Mitsuba |  |  |  |  |  |  |
| 6 | **Resident** | Mitsuba |  |  |  |  |  |  |
| 7 | **Resident** | Mitsuba |  |  |  |  |  |  |
| 8 | **Resident** | Mitsuba |  |  |  |  |  |  |
| 9 | **Resident** | Mitsuba |  |  |  |  |  |  |
| 10 | **Resident** | **Basil** |  |  |  |  |  |  |
| 11 | **Resident** | **Basil** |  |  |  |  |  |  |
| 12 | **Resident** | **Basil** |  |  |  |  |  |  |
| 13 | **Resident** | **Basil** |  |  |  |  |  |  |
| 14 | **Resident** | **Basil** |  |  |  |  |  |  |
| 15 | **Resident** | **Basil** |  |  |  |  |  |  |
| 16 | **Resident** | **Basil** |  |  |  |  |  |  |
| 17 | **Resident** | **Basil** |  |  |  |  |  |  |
| 18 | **Resident** | **Basil** |  |  |  |  |  |  |
| 19 | Staff | Mitsuba |  |  |  |  |  |  |
| 20 | Staff | Mitsuba |  |  |  |  |  |  |
| 21 | Staff | Mitsuba |  |  |  |  |  |  |
| 22 | Staff | Mitsuba |  |  |  |  |  |  |
| 23 | Staff | Mitsuba |  |  |  |  |  |  |
| 24 | Staff | Mitsuba |  |  |  |  |  |  |
| 25 | Staff | **Basil** |  |  |  |  |  |  |
| 26 | Staff | **Basil** |  |  |  |  |  |  |
| 27 | Staff | **Basil** |  |  |  |  |  |  |
| 28 | Staff | **Basil** |  |  |  |  |  |  |
| 29 | Staff | **Basil** |  |  |  |  |  |  |

**Q10. Within the designated area and during the designated period, please circle the ID numbers of any individuals who qualify as close contacts of the following resident. Responses are organised by date. The definitions of “close contact,” “designated area,” and “designated period” are the same as above.**

**Resident ID	5**

| **ID** | **Role** | **Unite** | **Wed, Dec 11**  **After 9:00 AM** | **Thu,Dec 12** | **Fri,Dec 13** | **Sat,Dec 14** | **Sun, Dec 15** | **Mon, Dec 16**  **Before 9:00AM** |
| --- | --- | --- | --- | --- | --- | --- | --- | --- |
| 1 | **Resident** | Mitsuba |  |  |  |  |  |  |
| 2 | **Resident** | Mitsuba |  |  |  |  |  |  |
| 3 | **Resident** | Mitsuba |  |  |  |  |  |  |
| 4 | **Resident** | Mitsuba |  |  |  |  |  |  |
| 5 | **Resident** | Mitsuba |  |  |  |  |  |  |
| 6 | **Resident** | Mitsuba |  |  |  |  |  |  |
| 7 | **Resident** | Mitsuba |  |  |  |  |  |  |
| 8 | **Resident** | Mitsuba |  |  |  |  |  |  |
| 9 | **Resident** | Mitsuba |  |  |  |  |  |  |
| 10 | **Resident** | **Basil** |  |  |  |  |  |  |
| 11 | **Resident** | **Basil** |  |  |  |  |  |  |
| 12 | **Resident** | **Basil** |  |  |  |  |  |  |
| 13 | **Resident** | **Basil** |  |  |  |  |  |  |
| 14 | **Resident** | **Basil** |  |  |  |  |  |  |
| 15 | **Resident** | **Basil** |  |  |  |  |  |  |
| 16 | **Resident** | **Basil** |  |  |  |  |  |  |
| 17 | **Resident** | **Basil** |  |  |  |  |  |  |
| 18 | **Resident** | **Basil** |  |  |  |  |  |  |
| 19 | Staff | Mitsuba |  |  |  |  |  |  |
| 20 | Staff | Mitsuba |  |  |  |  |  |  |
| 21 | Staff | Mitsuba |  |  |  |  |  |  |
| 22 | Staff | Mitsuba |  |  |  |  |  |  |
| 23 | Staff | Mitsuba |  |  |  |  |  |  |
| 24 | Staff | Mitsuba |  |  |  |  |  |  |
| 25 | Staff | **Basil** |  |  |  |  |  |  |
| 26 | Staff | **Basil** |  |  |  |  |  |  |
| 27 | Staff | **Basil** |  |  |  |  |  |  |
| 28 | Staff | **Basil** |  |  |  |  |  |  |
| 29 | Staff | **Basil** |  |  |  |  |  |  |

**Q11. Within the designated area and during the designated period, please circle the ID numbers of any individuals who qualify as close contacts of the following resident. Responses are organised by date. The definitions of “close contact,” “designated area,” and “designated period” are the same as above.**

**Resident ID	6**

| **ID** | **Role** | **Unite** | **Wed, Dec 11**  **After 9:00 AM** | **Thu,Dec 12** | **Fri,Dec 13** | **Sat,Dec 14** | **Sun, Dec 15** | **Mon, Dec 16**  **Before 9:00AM** |
| --- | --- | --- | --- | --- | --- | --- | --- | --- |
| 1 | **Resident** | Mitsuba |  |  |  |  |  |  |
| 2 | **Resident** | Mitsuba |  |  |  |  |  |  |
| 3 | **Resident** | Mitsuba |  |  |  |  |  |  |
| 4 | **Resident** | Mitsuba |  |  |  |  |  |  |
| 5 | **Resident** | Mitsuba |  |  |  |  |  |  |
| 6 | **Resident** | Mitsuba |  |  |  |  |  |  |
| 7 | **Resident** | Mitsuba |  |  |  |  |  |  |
| 8 | **Resident** | Mitsuba |  |  |  |  |  |  |
| 9 | **Resident** | Mitsuba |  |  |  |  |  |  |
| 10 | **Resident** | **Basil** |  |  |  |  |  |  |
| 11 | **Resident** | **Basil** |  |  |  |  |  |  |
| 12 | **Resident** | **Basil** |  |  |  |  |  |  |
| 13 | **Resident** | **Basil** |  |  |  |  |  |  |
| 14 | **Resident** | **Basil** |  |  |  |  |  |  |
| 15 | **Resident** | **Basil** |  |  |  |  |  |  |
| 16 | **Resident** | **Basil** |  |  |  |  |  |  |
| 17 | **Resident** | **Basil** |  |  |  |  |  |  |
| 18 | **Resident** | **Basil** |  |  |  |  |  |  |
| 19 | Staff | Mitsuba |  |  |  |  |  |  |
| 20 | Staff | Mitsuba |  |  |  |  |  |  |
| 21 | Staff | Mitsuba |  |  |  |  |  |  |
| 22 | Staff | Mitsuba |  |  |  |  |  |  |
| 23 | Staff | Mitsuba |  |  |  |  |  |  |
| 24 | Staff | Mitsuba |  |  |  |  |  |  |
| 25 | Staff | **Basil** |  |  |  |  |  |  |
| 26 | Staff | **Basil** |  |  |  |  |  |  |
| 27 | Staff | **Basil** |  |  |  |  |  |  |
| 28 | Staff | **Basil** |  |  |  |  |  |  |
| 29 | Staff | **Basil** |  |  |  |  |  |  |

**Q12. Within the designated area and during the designated period, please circle the ID numbers of any individuals who qualify as close contacts of the following resident. Responses are organised by date. The definitions of “close contact,” “designated area,” and “designated period” are the same as above.**

**Resident ID	7**

| **ID** | **Role** | **Unite** | **Wed, Dec 11**  **After 9:00 AM** | **Thu,Dec 12** | **Fri,Dec 13** | **Sat,Dec 14** | **Sun, Dec 15** | **Mon, Dec 16**  **Before 9:00AM** |
| --- | --- | --- | --- | --- | --- | --- | --- | --- |
| 1 | **Resident** | Mitsuba |  |  |  |  |  |  |
| 2 | **Resident** | Mitsuba |  |  |  |  |  |  |
| 3 | **Resident** | Mitsuba |  |  |  |  |  |  |
| 4 | **Resident** | Mitsuba |  |  |  |  |  |  |
| 5 | **Resident** | Mitsuba |  |  |  |  |  |  |
| 6 | **Resident** | Mitsuba |  |  |  |  |  |  |
| 7 | **Resident** | Mitsuba |  |  |  |  |  |  |
| 8 | **Resident** | Mitsuba |  |  |  |  |  |  |
| 9 | **Resident** | Mitsuba |  |  |  |  |  |  |
| 10 | **Resident** | **Basil** |  |  |  |  |  |  |
| 11 | **Resident** | **Basil** |  |  |  |  |  |  |
| 12 | **Resident** | **Basil** |  |  |  |  |  |  |
| 13 | **Resident** | **Basil** |  |  |  |  |  |  |
| 14 | **Resident** | **Basil** |  |  |  |  |  |  |
| 15 | **Resident** | **Basil** |  |  |  |  |  |  |
| 16 | **Resident** | **Basil** |  |  |  |  |  |  |
| 17 | **Resident** | **Basil** |  |  |  |  |  |  |
| 18 | **Resident** | **Basil** |  |  |  |  |  |  |
| 19 | Staff | Mitsuba |  |  |  |  |  |  |
| 20 | Staff | Mitsuba |  |  |  |  |  |  |
| 21 | Staff | Mitsuba |  |  |  |  |  |  |
| 22 | Staff | Mitsuba |  |  |  |  |  |  |
| 23 | Staff | Mitsuba |  |  |  |  |  |  |
| 24 | Staff | Mitsuba |  |  |  |  |  |  |
| 25 | Staff | **Basil** |  |  |  |  |  |  |
| 26 | Staff | **Basil** |  |  |  |  |  |  |
| 27 | Staff | **Basil** |  |  |  |  |  |  |
| 28 | Staff | **Basil** |  |  |  |  |  |  |
| 29 | Staff | **Basil** |  |  |  |  |  |  |

**Q13. Within the designated area and during the designated period, please circle the ID numbers of any individuals who qualify as close contacts of the following resident. Responses are organised by date. The definitions of “close contact,” “designated area,” and “designated period” are the same as above.**

**Resident ID	8**

| **ID** | **Role** | **Unite** | **Wed, Dec 11**  **After 9:00 AM** | **Thu,Dec 12** | **Fri,Dec 13** | **Sat,Dec 14** | **Sun, Dec 15** | **Mon, Dec 16**  **Before 9:00AM** |
| --- | --- | --- | --- | --- | --- | --- | --- | --- |
| 1 | **Resident** | Mitsuba |  |  |  |  |  |  |
| 2 | **Resident** | Mitsuba |  |  |  |  |  |  |
| 3 | **Resident** | Mitsuba |  |  |  |  |  |  |
| 4 | **Resident** | Mitsuba |  |  |  |  |  |  |
| 5 | **Resident** | Mitsuba |  |  |  |  |  |  |
| 6 | **Resident** | Mitsuba |  |  |  |  |  |  |
| 7 | **Resident** | Mitsuba |  |  |  |  |  |  |
| 8 | **Resident** | Mitsuba |  |  |  |  |  |  |
| 9 | **Resident** | Mitsuba |  |  |  |  |  |  |
| 10 | **Resident** | **Basil** |  |  |  |  |  |  |
| 11 | **Resident** | **Basil** |  |  |  |  |  |  |
| 12 | **Resident** | **Basil** |  |  |  |  |  |  |
| 13 | **Resident** | **Basil** |  |  |  |  |  |  |
| 14 | **Resident** | **Basil** |  |  |  |  |  |  |
| 15 | **Resident** | **Basil** |  |  |  |  |  |  |
| 16 | **Resident** | **Basil** |  |  |  |  |  |  |
| 17 | **Resident** | **Basil** |  |  |  |  |  |  |
| 18 | **Resident** | **Basil** |  |  |  |  |  |  |
| 19 | Staff | Mitsuba |  |  |  |  |  |  |
| 20 | Staff | Mitsuba |  |  |  |  |  |  |
| 21 | Staff | Mitsuba |  |  |  |  |  |  |
| 22 | Staff | Mitsuba |  |  |  |  |  |  |
| 23 | Staff | Mitsuba |  |  |  |  |  |  |
| 24 | Staff | Mitsuba |  |  |  |  |  |  |
| 25 | Staff | **Basil** |  |  |  |  |  |  |
| 26 | Staff | **Basil** |  |  |  |  |  |  |
| 27 | Staff | **Basil** |  |  |  |  |  |  |
| 28 | Staff | **Basil** |  |  |  |  |  |  |
| 29 | Staff | **Basil** |  |  |  |  |  |  |

**Q14. Within the designated area and during the designated period, please circle the ID numbers of any individuals who qualify as close contacts of the following resident. Responses are organised by date. The definitions of “close contact,” “designated area,” and “designated period” are the same as above.**

**Resident ID	9**

| **ID** | **Role** | **Unite** | **Wed, Dec 11**  **After 9:00 AM** | **Thu,Dec 12** | **Fri,Dec 13** | **Sat,Dec 14** | **Sun, Dec 15** | **Mon, Dec 16**  **Before 9:00AM** |
| --- | --- | --- | --- | --- | --- | --- | --- | --- |
| 1 | **Resident** | Mitsuba |  |  |  |  |  |  |
| 2 | **Resident** | Mitsuba |  |  |  |  |  |  |
| 3 | **Resident** | Mitsuba |  |  |  |  |  |  |
| 4 | **Resident** | Mitsuba |  |  |  |  |  |  |
| 5 | **Resident** | Mitsuba |  |  |  |  |  |  |
| 6 | **Resident** | Mitsuba |  |  |  |  |  |  |
| 7 | **Resident** | Mitsuba |  |  |  |  |  |  |
| 8 | **Resident** | Mitsuba |  |  |  |  |  |  |
| 9 | **Resident** | Mitsuba |  |  |  |  |  |  |
| 10 | **Resident** | **Basil** |  |  |  |  |  |  |
| 11 | **Resident** | **Basil** |  |  |  |  |  |  |
| 12 | **Resident** | **Basil** |  |  |  |  |  |  |
| 13 | **Resident** | **Basil** |  |  |  |  |  |  |
| 14 | **Resident** | **Basil** |  |  |  |  |  |  |
| 15 | **Resident** | **Basil** |  |  |  |  |  |  |
| 16 | **Resident** | **Basil** |  |  |  |  |  |  |
| 17 | **Resident** | **Basil** |  |  |  |  |  |  |
| 18 | **Resident** | **Basil** |  |  |  |  |  |  |
| 19 | Staff | Mitsuba |  |  |  |  |  |  |
| 20 | Staff | Mitsuba |  |  |  |  |  |  |
| 21 | Staff | Mitsuba |  |  |  |  |  |  |
| 22 | Staff | Mitsuba |  |  |  |  |  |  |
| 23 | Staff | Mitsuba |  |  |  |  |  |  |
| 24 | Staff | Mitsuba |  |  |  |  |  |  |
| 25 | Staff | **Basil** |  |  |  |  |  |  |
| 26 | Staff | **Basil** |  |  |  |  |  |  |
| 27 | Staff | **Basil** |  |  |  |  |  |  |
| 28 | Staff | **Basil** |  |  |  |  |  |  |
| 29 | Staff | **Basil** |  |  |  |  |  |  |

**Q15. Within the designated area and during the designated period, please circle the ID numbers of any individuals who qualify as close contacts of the following resident. Responses are organised by date. The definitions of “close contact,” “designated area,” and “designated period” are the same as above.**

**Resident ID	10**

| **ID** | **Role** | **Unite** | **Wed, Dec 11**  **After 9:00 AM** | **Thu,Dec 12** | **Fri,Dec 13** | **Sat,Dec 14** | **Sun, Dec 15** | **Mon, Dec 16**  **Before 9:00AM** |
| --- | --- | --- | --- | --- | --- | --- | --- | --- |
| 1 | **Resident** | Mitsuba |  |  |  |  |  |  |
| 2 | **Resident** | Mitsuba |  |  |  |  |  |  |
| 3 | **Resident** | Mitsuba |  |  |  |  |  |  |
| 4 | **Resident** | Mitsuba |  |  |  |  |  |  |
| 5 | **Resident** | Mitsuba |  |  |  |  |  |  |
| 6 | **Resident** | Mitsuba |  |  |  |  |  |  |
| 7 | **Resident** | Mitsuba |  |  |  |  |  |  |
| 8 | **Resident** | Mitsuba |  |  |  |  |  |  |
| 9 | **Resident** | Mitsuba |  |  |  |  |  |  |
| 10 | **Resident** | **Basil** |  |  |  |  |  |  |
| 11 | **Resident** | **Basil** |  |  |  |  |  |  |
| 12 | **Resident** | **Basil** |  |  |  |  |  |  |
| 13 | **Resident** | **Basil** |  |  |  |  |  |  |
| 14 | **Resident** | **Basil** |  |  |  |  |  |  |
| 15 | **Resident** | **Basil** |  |  |  |  |  |  |
| 16 | **Resident** | **Basil** |  |  |  |  |  |  |
| 17 | **Resident** | **Basil** |  |  |  |  |  |  |
| 18 | **Resident** | **Basil** |  |  |  |  |  |  |
| 19 | Staff | Mitsuba |  |  |  |  |  |  |
| 20 | Staff | Mitsuba |  |  |  |  |  |  |
| 21 | Staff | Mitsuba |  |  |  |  |  |  |
| 22 | Staff | Mitsuba |  |  |  |  |  |  |
| 23 | Staff | Mitsuba |  |  |  |  |  |  |
| 24 | Staff | Mitsuba |  |  |  |  |  |  |
| 25 | Staff | **Basil** |  |  |  |  |  |  |
| 26 | Staff | **Basil** |  |  |  |  |  |  |
| 27 | Staff | **Basil** |  |  |  |  |  |  |
| 28 | Staff | **Basil** |  |  |  |  |  |  |
| 29 | Staff | **Basil** |  |  |  |  |  |  |

**Q16. Within the designated area and during the designated period, please circle the ID numbers of any individuals who qualify as close contacts of the following resident. Responses are organised by date. The definitions of “close contact,” “designated area,” and “designated period” are the same as above.**

**Resident ID	11**

| **ID** | **Role** | **Unite** | **Wed, Dec 11**  **After 9:00 AM** | **Thu,Dec 12** | **Fri,Dec 13** | **Sat,Dec 14** | **Sun, Dec 15** | **Mon, Dec 16**  **Before 9:00AM** |
| --- | --- | --- | --- | --- | --- | --- | --- | --- |
| 1 | **Resident** | Mitsuba |  |  |  |  |  |  |
| 2 | **Resident** | Mitsuba |  |  |  |  |  |  |
| 3 | **Resident** | Mitsuba |  |  |  |  |  |  |
| 4 | **Resident** | Mitsuba |  |  |  |  |  |  |
| 5 | **Resident** | Mitsuba |  |  |  |  |  |  |
| 6 | **Resident** | Mitsuba |  |  |  |  |  |  |
| 7 | **Resident** | Mitsuba |  |  |  |  |  |  |
| 8 | **Resident** | Mitsuba |  |  |  |  |  |  |
| 9 | **Resident** | Mitsuba |  |  |  |  |  |  |
| 10 | **Resident** | **Basil** |  |  |  |  |  |  |
| 11 | **Resident** | **Basil** |  |  |  |  |  |  |
| 12 | **Resident** | **Basil** |  |  |  |  |  |  |
| 13 | **Resident** | **Basil** |  |  |  |  |  |  |
| 14 | **Resident** | **Basil** |  |  |  |  |  |  |
| 15 | **Resident** | **Basil** |  |  |  |  |  |  |
| 16 | **Resident** | **Basil** |  |  |  |  |  |  |
| 17 | **Resident** | **Basil** |  |  |  |  |  |  |
| 18 | **Resident** | **Basil** |  |  |  |  |  |  |
| 19 | Staff | Mitsuba |  |  |  |  |  |  |
| 20 | Staff | Mitsuba |  |  |  |  |  |  |
| 21 | Staff | Mitsuba |  |  |  |  |  |  |
| 22 | Staff | Mitsuba |  |  |  |  |  |  |
| 23 | Staff | Mitsuba |  |  |  |  |  |  |
| 24 | Staff | Mitsuba |  |  |  |  |  |  |
| 25 | Staff | **Basil** |  |  |  |  |  |  |
| 26 | Staff | **Basil** |  |  |  |  |  |  |
| 27 | Staff | **Basil** |  |  |  |  |  |  |
| 28 | Staff | **Basil** |  |  |  |  |  |  |
| 29 | Staff | **Basil** |  |  |  |  |  |  |

**Q17. Within the designated area and during the designated period, please circle the ID numbers of any individuals who qualify as close contacts of the following resident. Responses are organised by date. The definitions of “close contact,” “designated area,” and “designated period” are the same as above.**

**Resident ID	12**

| **ID** | **Role** | **Unite** | **Wed, Dec 11**  **After 9:00 AM** | **Thu,Dec 12** | **Fri,Dec 13** | **Sat,Dec 14** | **Sun, Dec 15** | **Mon, Dec 16**  **Before 9:00AM** |
| --- | --- | --- | --- | --- | --- | --- | --- | --- |
| 1 | **Resident** | Mitsuba |  |  |  |  |  |  |
| 2 | **Resident** | Mitsuba |  |  |  |  |  |  |
| 3 | **Resident** | Mitsuba |  |  |  |  |  |  |
| 4 | **Resident** | Mitsuba |  |  |  |  |  |  |
| 5 | **Resident** | Mitsuba |  |  |  |  |  |  |
| 6 | **Resident** | Mitsuba |  |  |  |  |  |  |
| 7 | **Resident** | Mitsuba |  |  |  |  |  |  |
| 8 | **Resident** | Mitsuba |  |  |  |  |  |  |
| 9 | **Resident** | Mitsuba |  |  |  |  |  |  |
| 10 | **Resident** | **Basil** |  |  |  |  |  |  |
| 11 | **Resident** | **Basil** |  |  |  |  |  |  |
| 12 | **Resident** | **Basil** |  |  |  |  |  |  |
| 13 | **Resident** | **Basil** |  |  |  |  |  |  |
| 14 | **Resident** | **Basil** |  |  |  |  |  |  |
| 15 | **Resident** | **Basil** |  |  |  |  |  |  |
| 16 | **Resident** | **Basil** |  |  |  |  |  |  |
| 17 | **Resident** | **Basil** |  |  |  |  |  |  |
| 18 | **Resident** | **Basil** |  |  |  |  |  |  |
| 19 | Staff | Mitsuba |  |  |  |  |  |  |
| 20 | Staff | Mitsuba |  |  |  |  |  |  |
| 21 | Staff | Mitsuba |  |  |  |  |  |  |
| 22 | Staff | Mitsuba |  |  |  |  |  |  |
| 23 | Staff | Mitsuba |  |  |  |  |  |  |
| 24 | Staff | Mitsuba |  |  |  |  |  |  |
| 25 | Staff | **Basil** |  |  |  |  |  |  |
| 26 | Staff | **Basil** |  |  |  |  |  |  |
| 27 | Staff | **Basil** |  |  |  |  |  |  |
| 28 | Staff | **Basil** |  |  |  |  |  |  |
| 29 | Staff | **Basil** |  |  |  |  |  |  |

**Q18. Within the designated area and during the designated period, please circle the ID numbers of any individuals who qualify as close contacts of the following resident. Responses are organised by date. The definitions of “close contact,” “designated area,” and “designated period” are the same as above.**

**Resident ID	13**

| **ID** | **Role** | **Unite** | **Wed, Dec 11**  **After 9:00 AM** | **Thu,Dec 12** | **Fri,Dec 13** | **Sat,Dec 14** | **Sun, Dec 15** | **Mon, Dec 16**  **Before 9:00AM** |
| --- | --- | --- | --- | --- | --- | --- | --- | --- |
| 1 | **Resident** | Mitsuba |  |  |  |  |  |  |
| 2 | **Resident** | Mitsuba |  |  |  |  |  |  |
| 3 | **Resident** | Mitsuba |  |  |  |  |  |  |
| 4 | **Resident** | Mitsuba |  |  |  |  |  |  |
| 5 | **Resident** | Mitsuba |  |  |  |  |  |  |
| 6 | **Resident** | Mitsuba |  |  |  |  |  |  |
| 7 | **Resident** | Mitsuba |  |  |  |  |  |  |
| 8 | **Resident** | Mitsuba |  |  |  |  |  |  |
| 9 | **Resident** | Mitsuba |  |  |  |  |  |  |
| 10 | **Resident** | **Basil** |  |  |  |  |  |  |
| 11 | **Resident** | **Basil** |  |  |  |  |  |  |
| 12 | **Resident** | **Basil** |  |  |  |  |  |  |
| 13 | **Resident** | **Basil** |  |  |  |  |  |  |
| 14 | **Resident** | **Basil** |  |  |  |  |  |  |
| 15 | **Resident** | **Basil** |  |  |  |  |  |  |
| 16 | **Resident** | **Basil** |  |  |  |  |  |  |
| 17 | **Resident** | **Basil** |  |  |  |  |  |  |
| 18 | **Resident** | **Basil** |  |  |  |  |  |  |
| 19 | Staff | Mitsuba |  |  |  |  |  |  |
| 20 | Staff | Mitsuba |  |  |  |  |  |  |
| 21 | Staff | Mitsuba |  |  |  |  |  |  |
| 22 | Staff | Mitsuba |  |  |  |  |  |  |
| 23 | Staff | Mitsuba |  |  |  |  |  |  |
| 24 | Staff | Mitsuba |  |  |  |  |  |  |
| 25 | Staff | **Basil** |  |  |  |  |  |  |
| 26 | Staff | **Basil** |  |  |  |  |  |  |
| 27 | Staff | **Basil** |  |  |  |  |  |  |
| 28 | Staff | **Basil** |  |  |  |  |  |  |
| 29 | Staff | **Basil** |  |  |  |  |  |  |

**Q19. Within the designated area and during the designated period, please circle the ID numbers of any individuals who qualify as close contacts of the following resident. Responses are organised by date. The definitions of “close contact,” “designated area,” and “designated period” are the same as above.**

**Resident ID	14**

| **ID** | **Role** | **Unite** | **Wed, Dec 11**  **After 9:00 AM** | **Thu,Dec 12** | **Fri,Dec 13** | **Sat,Dec 14** | **Sun, Dec 15** | **Mon, Dec 16**  **Before 9:00AM** |
| --- | --- | --- | --- | --- | --- | --- | --- | --- |
| 1 | **Resident** | Mitsuba |  |  |  |  |  |  |
| 2 | **Resident** | Mitsuba |  |  |  |  |  |  |
| 3 | **Resident** | Mitsuba |  |  |  |  |  |  |
| 4 | **Resident** | Mitsuba |  |  |  |  |  |  |
| 5 | **Resident** | Mitsuba |  |  |  |  |  |  |
| 6 | **Resident** | Mitsuba |  |  |  |  |  |  |
| 7 | **Resident** | Mitsuba |  |  |  |  |  |  |
| 8 | **Resident** | Mitsuba |  |  |  |  |  |  |
| 9 | **Resident** | Mitsuba |  |  |  |  |  |  |
| 10 | **Resident** | **Basil** |  |  |  |  |  |  |
| 11 | **Resident** | **Basil** |  |  |  |  |  |  |
| 12 | **Resident** | **Basil** |  |  |  |  |  |  |
| 13 | **Resident** | **Basil** |  |  |  |  |  |  |
| 14 | **Resident** | **Basil** |  |  |  |  |  |  |
| 15 | **Resident** | **Basil** |  |  |  |  |  |  |
| 16 | **Resident** | **Basil** |  |  |  |  |  |  |
| 17 | **Resident** | **Basil** |  |  |  |  |  |  |
| 18 | **Resident** | **Basil** |  |  |  |  |  |  |
| 19 | Staff | Mitsuba |  |  |  |  |  |  |
| 20 | Staff | Mitsuba |  |  |  |  |  |  |
| 21 | Staff | Mitsuba |  |  |  |  |  |  |
| 22 | Staff | Mitsuba |  |  |  |  |  |  |
| 23 | Staff | Mitsuba |  |  |  |  |  |  |
| 24 | Staff | Mitsuba |  |  |  |  |  |  |
| 25 | Staff | **Basil** |  |  |  |  |  |  |
| 26 | Staff | **Basil** |  |  |  |  |  |  |
| 27 | Staff | **Basil** |  |  |  |  |  |  |
| 28 | Staff | **Basil** |  |  |  |  |  |  |
| 29 | Staff | **Basil** |  |  |  |  |  |  |

**Q20. Within the designated area and during the designated period, please circle the ID numbers of any individuals who qualify as close contacts of the following resident. Responses are organised by date. The definitions of “close contact,” “designated area,” and “designated period” are the same as above.**

**Resident ID	15**

| **ID** | **Role** | **Unite** | **Wed, Dec 11**  **After 9:00 AM** | **Thu,Dec 12** | **Fri,Dec 13** | **Sat,Dec 14** | **Sun, Dec 15** | **Mon, Dec 16**  **Before 9:00AM** |
| --- | --- | --- | --- | --- | --- | --- | --- | --- |
| 1 | **Resident** | Mitsuba |  |  |  |  |  |  |
| 2 | **Resident** | Mitsuba |  |  |  |  |  |  |
| 3 | **Resident** | Mitsuba |  |  |  |  |  |  |
| 4 | **Resident** | Mitsuba |  |  |  |  |  |  |
| 5 | **Resident** | Mitsuba |  |  |  |  |  |  |
| 6 | **Resident** | Mitsuba |  |  |  |  |  |  |
| 7 | **Resident** | Mitsuba |  |  |  |  |  |  |
| 8 | **Resident** | Mitsuba |  |  |  |  |  |  |
| 9 | **Resident** | Mitsuba |  |  |  |  |  |  |
| 10 | **Resident** | **Basil** |  |  |  |  |  |  |
| 11 | **Resident** | **Basil** |  |  |  |  |  |  |
| 12 | **Resident** | **Basil** |  |  |  |  |  |  |
| 13 | **Resident** | **Basil** |  |  |  |  |  |  |
| 14 | **Resident** | **Basil** |  |  |  |  |  |  |
| 15 | **Resident** | **Basil** |  |  |  |  |  |  |
| 16 | **Resident** | **Basil** |  |  |  |  |  |  |
| 17 | **Resident** | **Basil** |  |  |  |  |  |  |
| 18 | **Resident** | **Basil** |  |  |  |  |  |  |
| 19 | Staff | Mitsuba |  |  |  |  |  |  |
| 20 | Staff | Mitsuba |  |  |  |  |  |  |
| 21 | Staff | Mitsuba |  |  |  |  |  |  |
| 22 | Staff | Mitsuba |  |  |  |  |  |  |
| 23 | Staff | Mitsuba |  |  |  |  |  |  |
| 24 | Staff | Mitsuba |  |  |  |  |  |  |
| 25 | Staff | **Basil** |  |  |  |  |  |  |
| 26 | Staff | **Basil** |  |  |  |  |  |  |
| 27 | Staff | **Basil** |  |  |  |  |  |  |
| 28 | Staff | **Basil** |  |  |  |  |  |  |
| 29 | Staff | **Basil** |  |  |  |  |  |  |

**Q21. Within the designated area and during the designated period, please circle the ID numbers of any individuals who qualify as close contacts of the following resident. Responses are organised by date. The definitions of “close contact,” “designated area,” and “designated period” are the same as above.**

**Resident ID	16**

| **ID** | **Role** | **Unite** | **Wed, Dec 11**  **After 9:00 AM** | **Thu,Dec 12** | **Fri,Dec 13** | **Sat,Dec 14** | **Sun, Dec 15** | **Mon, Dec 16**  **Before 9:00AM** |
| --- | --- | --- | --- | --- | --- | --- | --- | --- |
| 1 | **Resident** | Mitsuba |  |  |  |  |  |  |
| 2 | **Resident** | Mitsuba |  |  |  |  |  |  |
| 3 | **Resident** | Mitsuba |  |  |  |  |  |  |
| 4 | **Resident** | Mitsuba |  |  |  |  |  |  |
| 5 | **Resident** | Mitsuba |  |  |  |  |  |  |
| 6 | **Resident** | Mitsuba |  |  |  |  |  |  |
| 7 | **Resident** | Mitsuba |  |  |  |  |  |  |
| 8 | **Resident** | Mitsuba |  |  |  |  |  |  |
| 9 | **Resident** | Mitsuba |  |  |  |  |  |  |
| 10 | **Resident** | **Basil** |  |  |  |  |  |  |
| 11 | **Resident** | **Basil** |  |  |  |  |  |  |
| 12 | **Resident** | **Basil** |  |  |  |  |  |  |
| 13 | **Resident** | **Basil** |  |  |  |  |  |  |
| 14 | **Resident** | **Basil** |  |  |  |  |  |  |
| 15 | **Resident** | **Basil** |  |  |  |  |  |  |
| 16 | **Resident** | **Basil** |  |  |  |  |  |  |
| 17 | **Resident** | **Basil** |  |  |  |  |  |  |
| 18 | **Resident** | **Basil** |  |  |  |  |  |  |
| 19 | Staff | Mitsuba |  |  |  |  |  |  |
| 20 | Staff | Mitsuba |  |  |  |  |  |  |
| 21 | Staff | Mitsuba |  |  |  |  |  |  |
| 22 | Staff | Mitsuba |  |  |  |  |  |  |
| 23 | Staff | Mitsuba |  |  |  |  |  |  |
| 24 | Staff | Mitsuba |  |  |  |  |  |  |
| 25 | Staff | **Basil** |  |  |  |  |  |  |
| 26 | Staff | **Basil** |  |  |  |  |  |  |
| 27 | Staff | **Basil** |  |  |  |  |  |  |
| 28 | Staff | **Basil** |  |  |  |  |  |  |
| 29 | Staff | **Basil** |  |  |  |  |  |  |

**Q22. Within the designated area and during the designated period, please circle the ID numbers of any individuals who qualify as close contacts of the following resident. Responses are organised by date. The definitions of “close contact,” “designated area,” and “designated period” are the same as above.**

**Resident ID	17**

| **ID** | **Role** | **Unite** | **Wed, Dec 11**  **After 9:00 AM** | **Thu,Dec 12** | **Fri,Dec 13** | **Sat,Dec 14** | **Sun, Dec 15** | **Mon, Dec 16**  **Before 9:00AM** |
| --- | --- | --- | --- | --- | --- | --- | --- | --- |
| 1 | **Resident** | Mitsuba |  |  |  |  |  |  |
| 2 | **Resident** | Mitsuba |  |  |  |  |  |  |
| 3 | **Resident** | Mitsuba |  |  |  |  |  |  |
| 4 | **Resident** | Mitsuba |  |  |  |  |  |  |
| 5 | **Resident** | Mitsuba |  |  |  |  |  |  |
| 6 | **Resident** | Mitsuba |  |  |  |  |  |  |
| 7 | **Resident** | Mitsuba |  |  |  |  |  |  |
| 8 | **Resident** | Mitsuba |  |  |  |  |  |  |
| 9 | **Resident** | Mitsuba |  |  |  |  |  |  |
| 10 | **Resident** | **Basil** |  |  |  |  |  |  |
| 11 | **Resident** | **Basil** |  |  |  |  |  |  |
| 12 | **Resident** | **Basil** |  |  |  |  |  |  |
| 13 | **Resident** | **Basil** |  |  |  |  |  |  |
| 14 | **Resident** | **Basil** |  |  |  |  |  |  |
| 15 | **Resident** | **Basil** |  |  |  |  |  |  |
| 16 | **Resident** | **Basil** |  |  |  |  |  |  |
| 17 | **Resident** | **Basil** |  |  |  |  |  |  |
| 18 | **Resident** | **Basil** |  |  |  |  |  |  |
| 19 | Staff | Mitsuba |  |  |  |  |  |  |
| 20 | Staff | Mitsuba |  |  |  |  |  |  |
| 21 | Staff | Mitsuba |  |  |  |  |  |  |
| 22 | Staff | Mitsuba |  |  |  |  |  |  |
| 23 | Staff | Mitsuba |  |  |  |  |  |  |
| 24 | Staff | Mitsuba |  |  |  |  |  |  |
| 25 | Staff | **Basil** |  |  |  |  |  |  |
| 26 | Staff | **Basil** |  |  |  |  |  |  |
| 27 | Staff | **Basil** |  |  |  |  |  |  |
| 28 | Staff | **Basil** |  |  |  |  |  |  |
| 29 | Staff | **Basil** |  |  |  |  |  |  |

**Q23. Within the designated area and during the designated period, please circle the ID numbers of any individuals who qualify as close contacts of the following resident. Responses are organised by date. The definitions of “close contact,” “designated area,” and “designated period” are the same as above.**

**Resident ID	18**

| **ID** | **Role** | **Unite** | **Wed, Dec 11**  **After 9:00 AM** | **Thu,Dec 12** | **Fri,Dec 13** | **Sat,Dec 14** | **Sun, Dec 15** | **Mon, Dec 16**  **Before 9:00AM** |
| --- | --- | --- | --- | --- | --- | --- | --- | --- |
| 1 | **Resident** | Mitsuba |  |  |  |  |  |  |
| 2 | **Resident** | Mitsuba |  |  |  |  |  |  |
| 3 | **Resident** | Mitsuba |  |  |  |  |  |  |
| 4 | **Resident** | Mitsuba |  |  |  |  |  |  |
| 5 | **Resident** | Mitsuba |  |  |  |  |  |  |
| 6 | **Resident** | Mitsuba |  |  |  |  |  |  |
| 7 | **Resident** | Mitsuba |  |  |  |  |  |  |
| 8 | **Resident** | Mitsuba |  |  |  |  |  |  |
| 9 | **Resident** | Mitsuba |  |  |  |  |  |  |
| 10 | **Resident** | **Basil** |  |  |  |  |  |  |
| 11 | **Resident** | **Basil** |  |  |  |  |  |  |
| 12 | **Resident** | **Basil** |  |  |  |  |  |  |
| 13 | **Resident** | **Basil** |  |  |  |  |  |  |
| 14 | **Resident** | **Basil** |  |  |  |  |  |  |
| 15 | **Resident** | **Basil** |  |  |  |  |  |  |
| 16 | **Resident** | **Basil** |  |  |  |  |  |  |
| 17 | **Resident** | **Basil** |  |  |  |  |  |  |
| 18 | **Resident** | **Basil** |  |  |  |  |  |  |
| 19 | Staff | Mitsuba |  |  |  |  |  |  |
| 20 | Staff | Mitsuba |  |  |  |  |  |  |
| 21 | Staff | Mitsuba |  |  |  |  |  |  |
| 22 | Staff | Mitsuba |  |  |  |  |  |  |
| 23 | Staff | Mitsuba |  |  |  |  |  |  |
| 24 | Staff | Mitsuba |  |  |  |  |  |  |
| 25 | Staff | **Basil** |  |  |  |  |  |  |
| 26 | Staff | **Basil** |  |  |  |  |  |  |
| 27 | Staff | **Basil** |  |  |  |  |  |  |
| 28 | Staff | **Basil** |  |  |  |  |  |  |
| 29 | Staff | **Basil** |  |  |  |  |  |  |

**Q24. This question asks about your confidence in your responses. We will compare the close-contact list you selected in Q5, based on your own recollection, with the close-contact list identified from the device data. How well do you think these two lists will match? Please indicate your level of confidence by circling one option.**

1**. Not confident at all**

**2. Slightly confident**

**3. Neutral**

**4. Confident**

**5. Very Confident**

**Q25. This question asks about your confidence in your responses regarding residents. We will compare the resident close-contact list based on your recollection with the close-contact list identified from the device data. How well do you think these two lists will match? Please indicate your level of confidence by circling one option.**

1**. Not confident at all**

**2. Slightly confident**

**3. Neutral**

**4. Confident**

**5. Very Confident**

**Q26. Free comments (Please let us know if you have any feedback or suggestions for improving this questionnaire.)**

**This concludes with the questionnaire.**

**Before submitting, please check that you have not left any items unanswered.**

**Thank you very much for your cooperation.**
